# Effectiveness of the CoronaVac vaccine in the elderly population during a Gamma variant-associated epidemic of COVID-19 in Brazil: A test-negative case-control study

**DOI:** 10.1101/2021.05.19.21257472

**Authors:** Otavio T. Ranzani, Matt D.T. Hitchings, Murilo Dorion, Tatiana Lang D’Agostini, Regiane Cardoso de Paula, Olivia Ferreira Pereira de Paula, Edlaine Faria de Moura Villela, Mario Sergio Scaramuzzini Torres, Silvano Barbosa de Oliveira, Wade Schulz, Maria Almiron, Rodrigo Said, Roberto Dias de Oliveira, Patricia Vieira da Silva, Wildo Navegantes de Araújo, Jean Carlo Gorinchteyn, Jason R. Andrews, Derek A.T. Cummings, Albert I. Ko, Julio Croda

## Abstract

**Objective:** To estimate the effectiveness of the inactivated whole-virus vaccine, CoronaVac, against symptomatic COVID-19 in the elderly population of São Paulo State, Brazil during widespread circulation of the Gamma variant.

**Design:** Test negative case-control study.

**Setting:** Health-care facilities in São Paulo State, Brazil.

**Participants:** 43,774 adults aged 70 years or older who were residents of São Paulo State and underwent SARS-CoV-2 RT-PCR testing from January 17 to April 29, 2021. 26,433 cases with symptomatic COVID-19 and 17,622 symptomatic, test negative controls were selected into 7,950 matched pairs, according to age, sex, self-reported race, municipality of residence, prior COVID-19 status and date of RT-PCR testing.

**Intervention:** Vaccination with a two-dose regimen of CoronaVac.

**Main outcome measures:** RT-PCR confirmed symptomatic COVID-19 and COVID-19 associated hospitalizations and deaths.

**Results:** Adjusted vaccine effectiveness against symptomatic COVID-19 was 18.2% (95% CI, 0.0 to 33.2) in the period 0-13 days after the second dose and 41.6% (95% CI, 26.9 to 53.3) in the period ≥14 days after the second dose. Adjusted vaccine effectiveness against hospitalisations was 59.0% (95% CI, 44.2 to 69.8) and against deaths was 71.4% (95% CI, 53.7 to 82.3) in the period ≥14 days after the second dose. Vaccine effectiveness ≥14 days after the second dose declined with increasing age for the three outcomes, and among individuals aged 70-74 years it was 61.8% (95% CI, 34.8 to 77.7) against symptomatic disease, 80.1% (95% CI, 55.7 to 91.0) against hospitalisations and 86.0% (95% CI, 50.4 to 96.1) against deaths.

**Conclusions:** Vaccination with CoronaVac was associated with a reduction in symptomatic COVID-19, hospitalisations and deaths in adults aged 70 years or older in a setting with extensive Gamma variant transmission. However, significant protection was not observed until completion of the two-dose regimen, and vaccine effectiveness declined with increasing age amongst this elderly population.

**Summary boxes:** *What is already known on this topic:* Randomised controlled trials (RCT) have yielded varying estimates (51 to 84%) for the effectiveness of the inactivated whole-virus vaccine, CoronaVac, against symptomatic COVID-19. Current evidence is limited on whether CoronaVac is effective against severe disease or death caused by the SARS-CoV-2 variant of concern, Gamma, or in the setting of extensive Gamma variant circulation. More evidence is needed for the real-world effectiveness of CoronaVac and other inactivated vaccines among elderly individuals, a population that was underrepresented in RCTs of these vaccines.

*What this study adds:* A two-dose regimen of CoronaVac provides significant protection against symptomatic COVID-19, hospitalisations and deaths among adults ≥70 years of age in the setting of widespread Gamma variant transmission. Significant protection did not occur until ≥14 days after administration of the second dose of CoronaVac. The effectiveness of CoronaVac declines with increasing age in the elderly population.

## Introduction

The coronavirus disease (COVID-19) pandemic has caused 3.9 million deaths worldwide as of early July 2021,^1^ and has imparted disproportionately high mortality and morbidity on the elderly.^2^ A key question is whether the authorised COVID-19 vaccines are effective in the elderly, who may have impaired immune responses^3, 4^ and are underrepresented in randomised controlled trials (RCTs).^5–7^ mRNA and adenovirus vector-based vaccines have been shown to be effective against COVID-19 in elderly individuals,^8, 9^ but evidence is limited for the effectiveness of inactivated vaccines in these populations.^7, 10–12^

CoronaVac, an inactivated whole-virus vaccine, has been approved by 32 countries and jurisdictions,^10^ and has been implemented as part of mass vaccination campaigns in low-income and middle-income countries, many of which are experiencing COVID-19 epidemics due to the emergence of SARS-CoV-2 variants of concern (VOC). RCTs of a two-dose CoronaVac regimen in healthcare workers and the general population have yielded varying estimates (51 to 84%) of vaccine efficacy against symptomatic COVID-19.^5, 7, 10^ The World Health Organisation (WHO) Emergency Use Listing (EUL) procedure approved CoronaVac in early June 2021, but identified an evidence gap for the effectiveness of this vaccine in adults aged 60 and above.^11^ The WHO EUL cited an observational study in Chile,^10, 12^ which found that the adjusted effectiveness of CoronaVac, starting 14 days after the second dose, was 66.6% among adults aged 60 years and older. During the study period, the variant of concern (VOC) Gamma was detected in 28.6% of SARS-CoV-2 genomes.^12^ Furthermore, evidence from RCTs or observational studies have not addressed whether CoronaVac provides significant protection after administration of the first vaccine dose or in the setting of widespread VOC transmission.^5, 10, 11^

Brazil has experienced one of the world’s highest COVID-19 burdens during the pandemic with more than 18 million cases and 526,000 deaths as of early July 2021.^1, 13^ VOCs, and in particular the Gamma variant, have played an important role in the recent epidemic wave in Brazil which began in early 2021.^14–16^ The Gamma variant, which was first detected in Manaus, has increased transmissibility,^16^ has accrued mutations associated with decreased *in vitro* seroneutralisation,^17–19^ and at present, accounts for the majority of SARS-CoV-2 isolates genotyped in Brazil from 1 January 2021.^14, 20^ In the setting of a large Gamma variant-associated epidemic in São Paulo, the most populous state in Brazil, we conducted a matched, test-negative,^21^ case-control study to evaluate the real-world effectiveness of CoronaVac against symptomatic COVID-19 and severe clinical outcomes in the elderly population.

## Methods

### Study setting

The State of São Paulo (23°3ʹS, 46°4’W) has 645 municipalities and 46 million inhabitants, among which 3.23 million are ≥70 years of age.^22^ The state experienced three successive COVID-19 epidemic waves during which 2,997,282 cases (cumulative incidence rate: 6,475 per 100,000 population) and 100,649 deaths (cumulative mortality: 217 per 100,000 population) have been reported as of 9 May 2021 (Figure 1A, Supplementary Figure 1).^23^ The State Secretary of Health of Sao Paulo (SES-SP) initiated a COVID-19 vaccination campaign for the general population on 17 January 2021 according to an age-based prioritisation strategy (Figure 1, B-D) and is administering a two-dose regimen of CoronaVac, separated by a two to four week interval, and a two-dose regimen of ChAdOx1, separated by a 12 week interval.^24^ As of 29 April 2021, 8.63 million doses (5.16 first and 3.47 second million doses) have been administered of CoronaVac and 2.06 million doses (1.987 first and 0.07 second million doses) of ChAdOx1.

**Figure 1.**
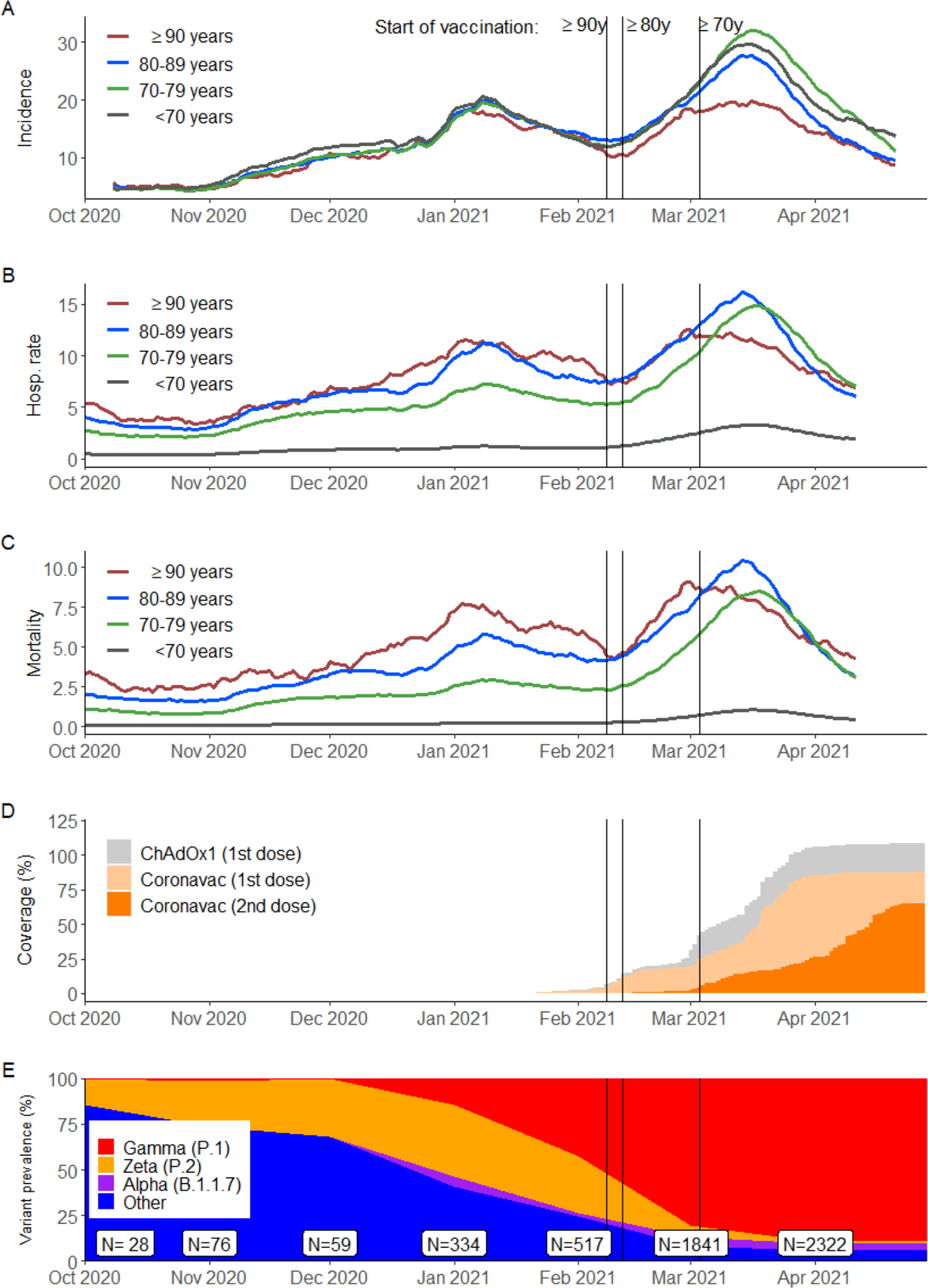
Incidence of reported COVID-19, vaccination coverage, and prevalence of SARS-CoV-2 variants of concern from Oct 1, 2020 to April 29, 2021 in São Paulo State, Brazil. Panels A, B, and C show the 14-day rolling average of daily age group-specific incidence of reported COVID-19 cases, hospitalization rate, and mortality (events per 100,000 population), respectively. Panel D shows daily cumulative vaccination coverage in individuals≥70 years of age. Population estimates for age groups were obtained from national projections for 2020.^20^ Panel E shows the monthly prevalence of SARS-CoV-2 variants among genotyped isolates in the GISAID database (extraction on June 20^th^ 2021).^18^ Vertical bars, from left to right in each panel, show the dates that adults ≥90, 80-89 and 70-79 years of age in the general population became eligible for vaccination.

### Study design

We conducted a matched test-negative case-control study to estimate the effectiveness of CoronaVac in reducing the odds of symptomatic RT-PCR-confirmed COVID-19 in adults ≥70 years of age from São Paulo State during the period from 17 January 2021, the start of COVID-19 vaccination, to 29 April 2021. Test-negative design studies have provided estimates of vaccine effectiveness in concordance with those obtained from RCTs^25, 26^ and have been used extensively to evaluate vaccines against respiratory infections,^27^ including COVID-19.^8, 21^ We chose the test-negative design because of the feasibility of accessing information on individuals who received SARS-CoV-2 testing from São Paulo State surveillance systems and the opportunity to control for potential biases, such as healthcare-seeking behaviour and access to testing.^21^ The study population was adults ≥70 years of age who had a residential address in São Paulo State, underwent SARS-CoV-2 RT-PCR testing during the study period, and had complete and consistent information between data sources on age, sex, residence, and vaccination and testing status and dates. We matched symptomatic test-negative controls to COVID-19 cases by date of testing to address potential sources of bias that may vary during the course of an epidemic, as well as by participant characteristics of age, gender, self-reported race, municipality of residence, and prior COVID-19 status.

The study design and statistical analysis plan were specified in advance of extracting information from data sources and are described in a publicly available protocol (https://github.com/juliocroda/VebraCOVID-19) and the Supplement. In the protocol, we pre-specified power thresholds for conducting analyses on the effectiveness of CoronaVac and ChAdOx1. These thresholds were achieved for CoronaVac but not for ChAdOx1 because of lower rates of ChAdOx1 administration in the population. We therefore restricted the evaluation of vaccine effectiveness to CoronaVac. The study was approved by the Ethical Committee for Research of Federal University of Mato Grosso do Sul (CAAE: 43289221.5.0000.0021).

### Data Sources

We obtained individual-level information on demographic characteristics, comorbidities, SARS-CoV-2 testing, and COVID-19 vaccination during the study period by extracting information on 6 May 2021 from the SES-SP laboratory testing registry (GAL), the national surveillance databases for COVID-19-like illnesses (e-SUS) and severe acute respiratory illness (SIVEP-Gripe), and the SES-SP vaccination registry (Vacina Já). Notification of suspected COVID-19 cases and SARS-CoV-2 testing results is compulsory in Brazil. The information technology bureau of the São Paulo State Government (PRODESP) linked individual-level records from the four databases using CPF numbers (Brazilian citizens’ unique identifier code) and provided anonymised datasets. We retrieved information on SARS-CoV-2 variants from genotyped isolates deposited in the GISAID database.^20^

### Selection of cases and matched controls

Cases were selected from the study population who had symptomatic COVID-19, defined as an individual who had a COVID-19-like illness; had a positive SARS-CoV-2 RT-PCR test result from a respiratory sample which was collected within 10 days after the onset of symptoms; and did not have a positive RT-PCR test in the preceding 90-day period. Controls were selected from the study population who had a COVID-19-like illness; had a negative SARS-CoV-2 RT-PCR test result from a respiratory sample that was collected within 10 days after the onset of symptoms;^21^ and did not have a positive RT-PCR test in the prior 90 days during the study period or in the subsequent 14 days. Cases and controls were excluded if they received the ChAdOx1 vaccine before sample collection for RT-PCR testing. COVID-19-like illness was defined as the presence of one or more reported COVID-19 related symptoms.^28^

We matched one test-negative control to each case according to RT-PCR sample collection date (±3 days); age category (5-year age bands, e.g, 70-74, 75-79 years); municipality of residence; self-reported race (defined as brown, black, yellow, white, or indigenous);^29^ and previous symptomatic events that were reported to the surveillance systems^28^ between February 1, 2020 and January 16, 2021, as a proxy for previous COVID-19 infection. Matching factors were chosen from variables that were associated with vaccination coverage or timing, and with SARS-CoV-2 infection risk or healthcare access (see protocol in Supplement).^21^ Upon identification of each case, a single control was randomly chosen from the set of all eligible matching controls.

### Statistical analysis

We estimated the effectiveness of CoronaVac against symptomatic COVID-19 during the periods 0-13 and ≥14 days after the second vaccine dose and ≥14 days after a single vaccine dose. Furthermore, we estimated the effectiveness of a single dose during the period 0-13 days after the first dose, when the vaccine has no or limited effectiveness.^5, 30, 31^ An association during this period may serve as an indicator of unmeasured confounding in the effectiveness estimate.^32^ The reference group for vaccination status was individuals who had not received a first vaccine dose before the date of sample collection.

We used conditional logistic regression to estimate the odds ratio (OR) of vaccination among cases and controls. 1-OR provided an estimate of vaccine effectiveness under the assumptions of a test-negative design.^33^ We included age and COVID-19-associated comorbidities (cardiovascular, renal, neurological, haematological, or hepatic comorbidities, diabetes, chronic respiratory disorder, obesity, or immunosuppression) as covariates in the model. We evaluated nonlinearity for age using restricted cubic splines and chose the parsimonious model comparing nested models with a likelihood ratio test. Furthermore, we conducted a *post hoc* sensitivity analysis that incorporated the calendar date of RT-PCR sample collection in the model to evaluate potential residual confounding that may not be addressed by the matching criteria We estimated the vaccine effectiveness against acute respiratory illness (ARI) associated hospitalizations and deaths in a *post hoc* analysis. In separate analyses, we selected matched pairs in which the case had the secondary outcome of interest.^34, 35^ We fit the same conditional logistic regression model as for the primary outcome.

We conducted a pre-specified analysis of vaccine effectiveness among age sub-groups for the primary and secondary outcomes, but could not perform analyses stratified by previous COVID-19 documented infection because of small numbers. Additional *post hoc* analyses were performed of vaccine effectiveness for the primary outcome for subgroups stratified by sex, number of chronic comorbidities (none vs. at least one), the two most frequent chronic comorbidities (cardiovascular disease and diabetes), and region of residence (“Grande São Paulo” health region vs. others). Interaction terms were incorporated into the model to evaluate the association of each subgroup of interest with vaccine effectiveness ≥14 days after the second dose.

### Power calculation

Our protocol specified that we would conduct proposed analyses after achieving ≥80% power to identify a vaccine effectiveness of 40% against symptomatic COVID-19 for the comparison of ≥14 days after the second dose of CoronaVac and not receiving a vaccine dose. The power was simulated fitting conditional logistic regressions on 1,000 simulated datasets. After extracting the surveillance databases on May 6, 2021 and generating matched case-control pairs, we determined that the power of the study was 99.9% and proceeded to conduct the pre-specified analyses. We did not perform an analysis for ChAdOx1 since the simulated power was 31% to identify a vaccine effectiveness of 40% for the comparison of ≥28 days after the first dose of ChAdOx1 and not receiving a vaccine dose. All analyses were done in R, version 4.0.2.

## Results

### COVID-19 epidemic and vaccination campaign in São Paulo State

São Paulo State experienced three COVID-19 epidemic waves during which peak incidence occurred in July 2020 for the first wave (Supplementary Figure 1), January 2021 for the second wave and March 2021 for the third wave (Figure 1A). The second wave was preceded in November 2020 by an increase in the prevalence of the Zeta variant among genotyped isolates from São Paulo State deposited into the GISAID database (Figure 1E). The third wave was preceded in January 2021 by an increase in the prevalence of the Gamma variant among genotyped isolates. The Gamma variant replaced other SARS-CoV-2 variants^20^ and accounted for 79% (3,834/4,887) of the genotyped isolates that were reported in GISAID during the study period and 86% (3,584/4,192) of genotyped isolates that were reported between 1 March to 29 April 2021 when the majority of discordant case-control pairs were identified (Supplementary Figure 2). The vaccination campaign, initiated on January 17, 2021, achieved an estimated coverage of roughly 85% for the first (2.82 million) and 65% for second (2.10 million) CoronaVac doses among adults ≥70 years of age by April 29, 2021 (Figure 1B-D). After initiation of the vaccination campaign and during the third epidemic wave, COVID-19 incidence increased and peaked in late March in all age groups except for adults ≥90 years of age (Figure 1A).

### Study population

Among 43,774 individuals eligible for study inclusion (Figure 2), 15,852 (36.2%) who provided 15,900 RT-PCR test results were selected into 7,950 matched case and control pairs. There were 38 individuals that contributed two times as controls and 10 individuals one time as control and one time as case. Table 1 shows the characteristics of eligible individuals with positive and negative RT-PCR tests and selected cases and matched controls. A higher proportion of cases had reported comorbidities than controls. Supplementary Table 1 shows the distribution of matched pairs according to the vaccination status of cases and controls at the time of RT-PCR testing. The majority of discordant pairs, based on vaccination status, were selected after 14 March 2021 (Supplementary Figure 3). Cases and controls who completed the two dose vaccine regimen had similar inter-dose intervals (mean 29 vs. 25 days). Likewise, cases and controls who were vaccinated had similar distributions for the intervals between administration of vaccine doses and RT-PCR testing (Table 1 and Supplementary Figure 3). The characteristics of the matched case and control pairs which were selected for the analysis of secondary outcomes of hospitalisation (n=8,078) and death (n=4,104) are shown in Supplementary Tables 2 and 3.

**Figure 2.**
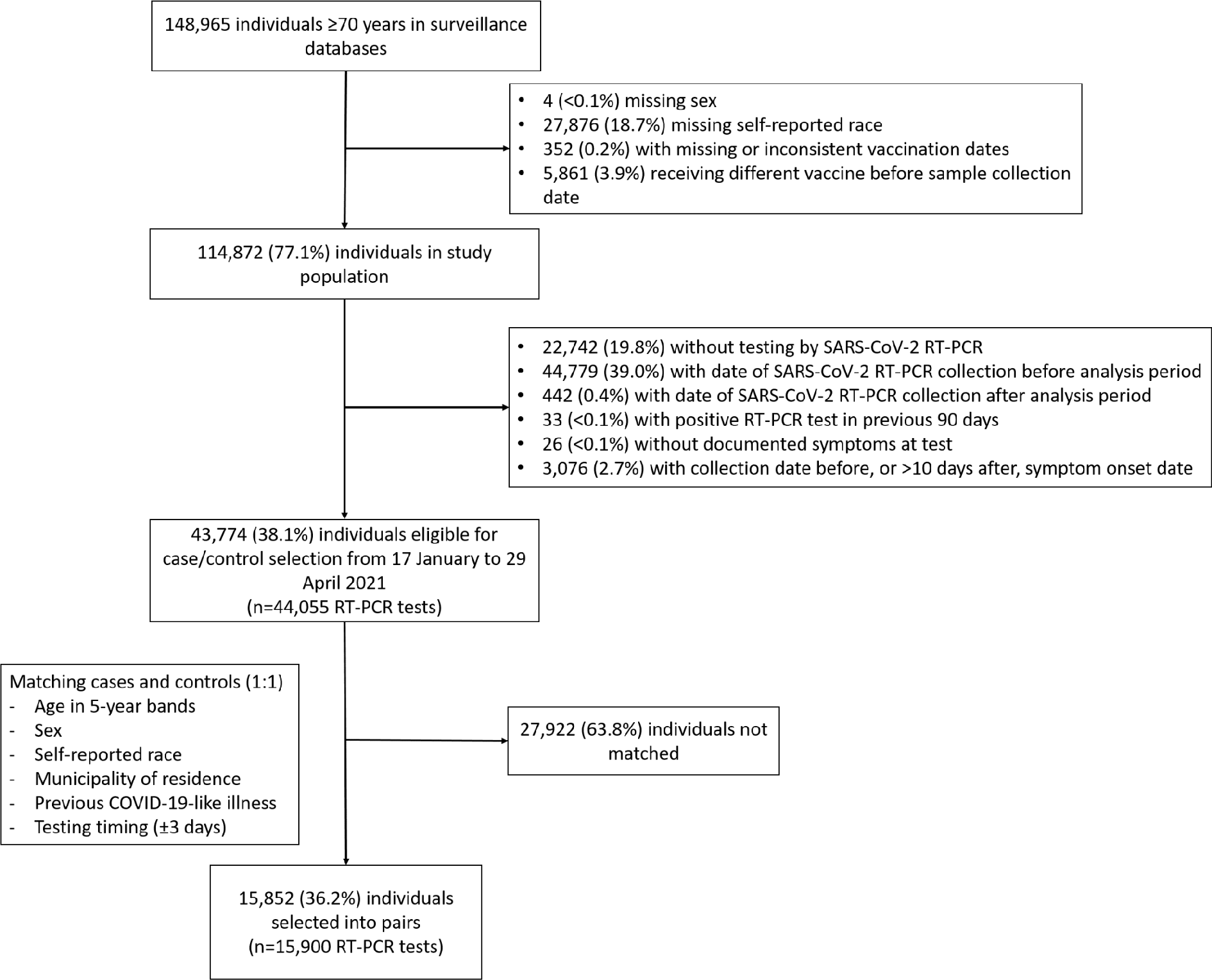
Flowchart of the identification of the study population from surveillance databases and selection of matched cases and controls.

**Figure 3.**
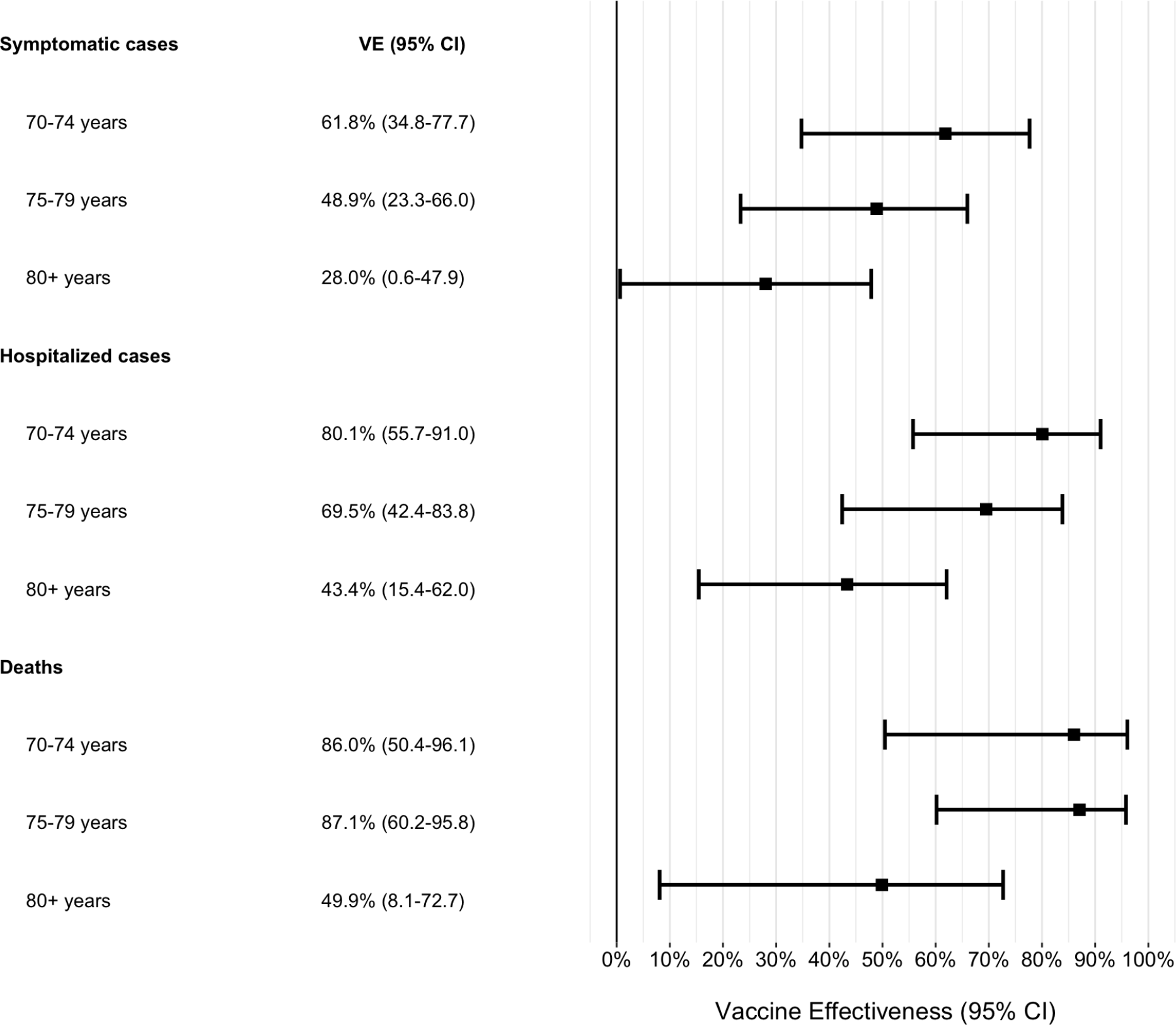
Adjusted vaccine effectiveness during the period ≥14 days after the second CoronaVac dose for subgroups of adults ≥70 years of age. Estimates of vaccine effectiveness were obtained from a conditional logistic regression model that included covariates of age and the number of comorbidities and incorporated an interaction term between the category of interest and the period ≥14 days after the second CoronaVac dose.

**Table 1.** Characteristics of adults ≥70 years of age who were eligible for matching and selected into case-test negative pairs.

| Characteristics* | Eligible cases and controls |  | Matched pairs |  |
| --- | --- | --- | --- | --- |
|  | Test-negative<br>(n=17,622)^ | Test-positive<br>(n=26,433)^ | Controls<br>(n=7,950)^ | Cases<br>(n=7,950)^ |
| **Demographics** |  |  |  |  |
| Age, mean (SD), years | 77.53 (6.8) | 76.71 (6.2) | 76.15 (5.8) | 76.15 (5.8) |
| Age categories, n (%) |  |  |  |  |
| 70-79 years | 12,123 (68.8) | 19,673 (74.4) | 6,150 (77.4) | 6,150 (77.4) |
| 80-89 years | 4,301 (24.4) | 5,437 (20.6) | 1,510 (19.0) | 1,510 (19.0) |
| $\geq 90$ years | 1,198 (6.8) | 1,323 (5.0) | 290 (3.6) | 290 (3.6) |
| Male sex, n (%) | 7,689 (43.6) | 12,431 (47.0) | 3,276 (41.2) | 3,276 (41.2) |
| Self-reported race<sup>†</sup>, n (%) |  |  |  |  |
| White/Branca | 13,415 (76.1) | 19,796 (74.9) | 6,420 (80.8) | 6,420 (80.8) |
| Brown/Pardo | 3,192 (18.1) | 4,983 (18.9) | 1,301 (16.4) | 1,301 (16.4) |
| Black/Preta | 785 (4.5) | 1,258 (4.8) | 191 (2.4) | 191 (2.4) |
| Yellow/ Amarela | 226 (1.3) | 390 (1.5) | 38 (0.5) | 38 (0.5) |
| Indigenous/Indigena | 4 (0.0) | 6 (0.0) | - | - |
| Residence in “Grande São Paulo”<br>Health Region, n (%) | 12,381 (70.3) | 16,538 (62.6) | 4,259 (53.6) | 4,259 (53.6) |
| **Comorbidities** |  |  |  |  |
| Reported number<sup>‡</sup>, n (%) |  |  |  |  |
| None | 10,027 (56.9) | 12,668 (47.9) | 4,510 (56.7) | 3,564 (44.8) |
| One or two | 6,984 (39.6) | 12,548 (47.5) | 3,151 (39.6) | 3,994 (50.2) |
| Three or more | 611 (3.5) | 1,217 (4.6) | 289 (3.6) | 392 (4.9) |
| Cardiovascular disease , n (%) | 5,293 (30.0) | 10,079 (38.1) | 2,375 (29.9) | 3,252 (40.9) |
| Diabetes, n (%) | 3,233 (18.3) | 6,533 (24.7) | 1,314 (19.0) | 2,092 (26.3) |
| **Prior SARS-CoV-2 exposure \*\*** |  |  |  |  |
| Previous symptomatic events notified to the surveillance systems**, n (%) | 685 (3.9) | 354 (1.3) | 35 (0.4) | 35 (0.4) |
| Positive SARS-CoV-2 test result <sup>††</sup>, n (%) | 66 (0.4) | 13 (0.0) | 1 (0.0) | 4 (0.1) |
| Interval between symptoms onset and RT-PCR testing, median (p25-p75), days | 3 [2-5] | 4 [2-6] | 3 [1-5] | 4 [2-6] |
| **ARI associated hospitalisations, n (%)** | 4,524/17,484 (25.9) | 12,987/26,221 (49.5) | 2,065/7,889 (26.2) | 4,039/7,883 (51.2) |
| **ARI associated deaths, n (%)** | 912/16,710 (5.5%) | 7,054/24,508 (28.8%) | 729/7,557 (9.6%) | 2,052/7,359 (27.9%) |
| Interval between symptoms onset and hospitalization, median (p25-p75), days | 3 [2-6] | 7 [4-10] | 3 [2-6] | 7 [4-10] |
| Interval between symptoms onset and deaths, median (p25-p75), days | 8 [4-13] | 14 [9-21] | 8 [4-15] | 15 [10-22] |
| **Vaccination status** |  |  |  |  |
| Not vaccinated, n (%) | 11,986 (68.0) | 17,233 (65.2) | 5,485 (69.0) | 5,561 (69.9) |
| Single dose, within 0-13 days, n (%) | 1,446 (8.2) | 2,976 (11.3) | 747 (9.4) | 762 (9.6) |
| Single dose, ≥14 days, n (%) | 1,797 (10.2) | 3,312 (12.5) | 843 (10.6) | 851 (10.7) |
| Two doses, within 0-13 days, n (%) | 1,041 (5.9) | 1,533 (5.8) | 437 (5.5) | 421 (5.3) |
| Two doses, ≥14 days, n (%) | 1,352 (7.7) | 1,379 (5.2) | 438 (5.5) | 355 (4.5) |
| Interval between first and second dose, mean (SD), days | 25 (6) | 30 (12) | 25 (6) | 29 (11) |
| Interval between first dose and RT-PCR testing, mean (SD), days | 28 (19) | 23 (16) | 24 (17) | 23 (16) |
| Interval between second dose and RT-PCR testing, mean (SD), days | 20 (15) | 17 (14) | 18 (15) | 17 (14) |

\*Continuous variables are displayed as mean (SD); categorical variables are displayed as n (%).
^These numbers refer to RT-PCR tests and represent 43,774 individuals for the eligible cases and controls and 15,852 individuals in the matched cases and controls.
<sup>†</sup>Race/skin colour as defined by the Brazilian national census bureau (Instituto Nacional de Geografia e Estatísticas).<sup>27</sup>
<sup>‡</sup>Comorbidities included: cardiovascular, renal, neurological, haematological, or hepatic comorbidities, diabetes, chronic respiratory disorder, obesity, or immunosuppression.
\*\* Prior to the start of the study on 17 January, 2021 and after systematic surveillance was implemented on 1 February, 2020.
\*\* Reported illness with COVID-19 associated symptoms in the eSUS and SIVEP-Gripe databases.
<sup>††</sup> Defined as a positive SARS-CoV-2 RT-PCR or antigen detection test result.

**Table 2:** Effectiveness of CoronaVac against symptomatic COVID-19, hospitalisations and deaths in adults ≥70 years of age.

|  | Unadjusted Analysis |  |  | Adjusted Analysis<sup>^</sup> |  |  |
| --- | --- | --- | --- | --- | --- | --- |
| Symptomatic COVID-19 (n=15,900) | OR (95% CI) | VE (95% CI) | p-value | OR (95% CI) | VE (95% CI) | p-value |
| Single dose, within 0-13 days vs. unvaccinated* | 0.97 (0.85-1.12) | 2.7% (-11.7-15.3) | 0.70 | 0.98 (0.85-1.12) | 2.5% (-12.2-15.3) | 0.72 |
| Single dose, ≥14 days vs. unvaccinated* | 0.91 (0.78-1.05) | 9.5% (-5.3-22.3) | 0.20 | 0.90 (0.77-1.04) | 10.5% (-4.4-23.3) | 0.16 |
| Two doses, within 0-13 days vs. unvaccinated* | 0.81 (0.66-0.98) | 19.5% (1.9-34.0) | 0.03 | 0.82 (0.67-1.00) | 18.2% (0.0-33.2) | 0.05 |
| Two doses, ≥14 days vs. unvaccinated* | 0.60 (0.48-0.74) | 40.5% (25.8-52.3) | <0.001 | 0.58 (0.47-0.73) | 41.6% (26.9-53.3) | <0.001 |
| **COVID-19 associated hospitalisations (n=8,078)** |  |  |  |  |  |  |
| Single dose, within 0-13 days vs. unvaccinated* | 0.89 (0.74-1.07) | 11.3% (-7.0-26.4) | 0.21 | 0.84 (0.68-1.02) | 16.4% (-2.2-31.6) | 0.08 |
| Single dose, ≥14 days vs. unvaccinated* | 0.85 (0.70-1.04) | 14.6% (-4.2-30.0) | 0.12 | 0.83 (0.66-1.01) | 18.5% (-1.0-34.2) | 0.06 |
| Two doses, within 0-13 days vs. unvaccinated* | 0.62 (0.47-0.81) | 38.1% (18.8-52.8) | 0.001 | 0.59 (0.44-0.79) | 40.9% (20.7-55.9) | <0.001 |
| Two doses, ≥14 days vs. unvaccinated* | 0.47 (0.36-0.63) | 52.7% (37.2-64.4) | <0.001 | 0.41 (0.30-0.56) | 59% (44.2-69.8) | <0.001 |
| **COVID-19 associated deaths (n=4,104)** |  |  |  |  |  |  |
| Single dose, within 0-13 days vs. unvaccinated* | 0.92 (0.72-1.18) | 8.2% (-17.7-28.4) | 0.50 | 0.93 (0.71-1.21) | 7.4% (-21.3-29.2) | 0.58 |
| Single dose, ≥14 days vs. unvaccinated* | 0.76 (0.57-1.00) | 24.5% (0.0-43.0) | 0.05 | 0.68 (0.50-0.93) | 31.6% (7.1-49.7) | 0.02 |
| Two doses, within 0-13 days vs. unvaccinated* | 0.40 (0.27-0.59) | 60.4% (40.6-73.5) | <0.001 | 0.36 (0.23-0.55) | 64.4% (44.6-77.1) | <0.001 |
| Two doses, ≥14 days vs. unvaccinated* | 0.34 (0.22-0.52) | 66.2% (47.8-78.1) | <0.001 | 0.29 (0.18-0.46) | 71.4% (53.7-82.3) | <0.001 |

ARI - acute respiratory illness
\*At date of index sample collection for cases and controls.
<sup>^</sup> Models adjusted by age (linear term for symptomatic and hospitalisation, restricted cubic spline for deaths) and number of comorbidities (None, One or Two, Three or more)

### Vaccine effectiveness

The adjusted effectiveness of the two-dose CoronaVac schedule against symptomatic COVID-19 was 18.2% (95% CI 0.0 to 33.2) in the period 0-13 days and 41.6% (95% CI 26.9 to 53.3) in the period ≥14 days after administration of the second dose (Table 2). We did not identify a significant reduction or increase in the odds of COVID-19 in the time periods following a single vaccine dose, including the period 0-13 days which serves as a potential bias-indicator. Increasing number of comorbidities was significantly associated with increased odds of COVID-19. In a sensitivity analysis including calendar date of testing as a covariate, vaccine effectiveness was 19.3% (95% CI 1.3 to 34) in the period 0-13 day and 42.3% (95% CI 27.7 to 53.9) in the period ≥14 days after administration of the second dose.

In the period starting 14 days after the second dose, the adjusted effectiveness of the two-dose schedule was 59.0% (95% CI 44.2 to 69.8) against hospitalisation and 71.4% (95% CI 53.7 to 82.3) against deaths (Table 2). In general, statistically significant protection was not observed until after the second dose, and the vaccine effectiveness in the “bias-indicator” period 0-13 days after the first dose was low.

Vaccine effectiveness against symptomatic COVID-19 in the period ≥14 days after the second dose declined with increasing age and was 61.8% (95% CI 34.8 to 77.7) among individuals 70-74 years old, 48.9% (95% CI 23.3 to 66.0) among 75-79 years old, and 28.0% (95% CI 0.6 to 47.9) among individuals ≥80 years of age (p_interaction_ = 0.05)(Figure 3). The same pattern was observed for hospitalisations (p_interaction_ = 0.04) and deaths (p_interaction_ = 0.19), yielding effectiveness of 80.1% (95% CI 55.7 to 91.0) for hospitalisations and 86.0% (95% CI 34.8 to 77.7) for deaths among the 70-74 years age group (Figure 3 and Supplementary Table 4).

Vaccine effectiveness against symptomatic COVID-19 disease did not differ among sub-groups defined by sex, presence of comorbidities, reported cardiovascular disease, or regions of residence. However, individuals with reported diabetes had lower protection than those without reported diabetes (VE 26.9% vs. 45.6%, p_interaction_ = 0.12) during the period starting 14 days after the 2nd dose (Supplementary Table 5 and Supplementary Figure 4).

## Discussion

This test-negative case-control study found that a two-dose schedule of CoronaVac had a real-world effectiveness of 41.6% (95% CI 26.9 to 53.3) against symptomatic COVID-19, 59.0% (95% CI 44.2 to 69.8) against COVID associated hospitalisations, and 71.4% (95% CI 53.7 to 82.3%) against COVID-19 associated deaths among those ≥70 years during a Gamma variant-associated epidemic in Brazil. Furthermore, we have addressed several evidence gaps for the use of this vaccine: 1) vaccination with CoronaVac demonstrated an effectiveness against COVID-19, including associated severe outcomes, in the setting of widespread Gamma transmission which was similar to that found in the Brazilian RCT conducted prior to the emergence of Gamma,^5^ 2) the vaccine did not confer significant protection until 14 days after completion of the two dose regimen; and 3) vaccine effectiveness declined with increasing age among adults ≥70 years of age.

### Research in context

A key evidence gap, as raised in the WHO EUL for Coronavac,^11^ has been the effectiveness of this vaccine in the elderly population, since this age group was not represented in the Brazilian and Turkish RCTs.^5, 7, 10, 11^ We found that CoronaVac had an effectiveness in the elderly population that was similar to that observed in RCTs of younger populations and similar to estimates of vaccine effectiveness in adults ≥60 years of age from a retrospective cohort study in Chile.^10, 12^ However, we observed a significant decline in vaccine effectiveness against symptomatic COVID-19 with increasing age from 61.8% (95% CI 34.8 to 77.7) in adults 70-74 year olds to 28.0% (95% CI 0.6 to 47.9) in adults ≥80 years of age. These findings parallel real-world evidence for the BNT162b2 mRNA vaccine, which found reduced effectiveness in residents of long-term care facilities in Denmark,^36^ skilled nursing facilities in the USA,^37^ and the general population with ≥70 years in Finland^38^ and ≥80 years of age in Israel.^39^ As well as a slower immune response and lower peak of neutralising antibodies than younger populations, elderly individuals seem to have faster decay of antibodies titers.^4^ Together, these findings suggest that effective COVID-19 vaccination of the very elderly (≥80 years) population may require specific vaccines or vaccination schemes.

Vaccine effectiveness was greater against severe outcomes than against symptomatic COVID-19 in all age subgroups among the elderly. This finding, consistent with RCTs and observational studies for multiple COVID-19 vaccines and across settings,^5, 6, 9, 10, 12^ suggests that vaccination will reduce morbidity and mortality even if effectiveness at preventing infections is reduced among the elderly. The direct comparison of the effectiveness against hospitalisation with other vaccines and between countries is not straightforward, because hospitalisation is dependent on admission triage policies that change according to age and hospital bed availability. Therefore, a patient above 80 years with symptomatic COVID-19 has higher likelihood of being admitted compared to younger patients even if not severe, and this likelihood varies between public and private facilities and whether the health system is overwhelmed.^13^ Thus, we cannot generalise our findings for protection against hospitalisations without considering this context. We evaluated vaccine effectiveness at the individual level, not accounting for the indirect effect and the total effect from the vaccination campaign. A preliminary aggregated analysis using weekly times series of COVID-19 deaths in Brazil found a relative decrease in mortality among those ≥70 years compared with all ages after the vaccination with CoronaVac and ChAdOx1,^40^ suggesting a discernible impact of vaccination on mortality at the population level. Additional investigation is required to address the duration of protection conferred by Coronavac.^7, 19, 21^

The absence of demonstrable effectiveness of CoronaVac until completion of the two dose regimen has profound implications for its use in an epidemic response. In contrast to COVID-19 vaccines that confer protection after the first dose,^9, 41^ we did not detect significant effectiveness for CoronaVac until ≥14 days after the second dose (more than six weeks after the first dose).^19^ Our findings suggest that in countries where CoronaVac supplies are constrained and are experiencing high SARS-CoV-2 transmission, vaccination should prioritise completion of the two-dose regimen among the highest risk populations and avoid expanding to broader segments for which provisions for a second dose have not been secured.

Our study did not directly address the question whether vaccination with CoronaVac was effective against Gamma-variant-associated COVID-19 since we have no data on whether the analysed cases were due to Gamma variant. However, 90% (1,790/1,999) of the discordant pairs in this matched case-control study were selected during the period 1 March to 29 April 2021, when Gamma accounted for 85% of the genotyped isolates during surveillance in São Paulo state. A test-negative study in Canada evaluated ≥70 years individuals and estimated an adjusted vaccine effectiveness of single-dose mRNA vaccines of 61% (95% CI 45-72) against the VOC Gamma compared to 72% (95% CI 58-81) for non-VOC.^42^ Although further studies are required to determine the effectiveness of CoronaVac against Gamma and additional VOCs, our findings provide supportive evidence for the use of CoronaVac in countries in South America which are experiencing epidemics due to extensive spread of Gamma^20^ and are administering mass vaccination with CoronaVac as part of the epidemic response.

### Strengths and limitations of this study

This study has several strengths which include the large sample size and geospatial coverage, comprising the state of São Paulo with 46 million inhabitants distributed across 645 municipalities. We implemented a pre-specified publicly-available protocol, which is in accordance with the recent WHO guideline for COVID-19 vaccine effectiveness evaluation.^21^ Using a test-negative design, we have addressed biases that affect observational vaccine effectiveness studies, such as health-seeking behaviour and access. Additionally, after matching and adjustment, the “bias-indicator” association between recent vaccination with a single dose 0-13 days before sample collection was close to null, suggesting that vaccinated and unvaccinated individuals did not differ in their underlying risk of testing positive for SARS-CoV-_2_^.8, 32, 43^

Our study had limitations. We could not assess the influence of a previous SARS-CoV-2 infection on vaccine effectiveness since passive surveillance identified few individuals with a positive RT-PCR or rapid antigen test before the study period. Prior to the start of the vaccination campaign, the estimated seroprevalence of COVID-19 in inhabitants who were ≥60 years of age in the capital of São Paulo State was 19.9% (95% CI, 14.9-29.9) in January 2021.^44^ Our estimates of vaccine effectiveness may therefore be subject to downward bias as unvaccinated individuals were at lower risk of reinfection. We attempted to exclude false-negative RT-PCR tests by excluding as controls patients with a subsequent positive test within 14 days after the initial testing and including only tests performed 10 days of symptom onset.^21^ In addition, we restricted our study population to elderly individuals because they were a priority group for vaccination and received the large majority of CoronaVac doses during the initial stages of the campaign in Brazil; as a result, a direct comparison of the effectiveness of CoronaVac between older and younger populations was not possible. Our analyses were also limited by the lack of more refined covariates, such as frailty and chronic illness status, which could influence vaccine effectiveness in the very elderly and would not be addressed by age and reported comorbidities *per se*. Finally, although we matched for calendar time of SARS-CoV-2 testing (±3 days),^21^ we cannot exclude the possibility of time-varying changes in behaviour or testing practices among participants that were not addressed by our matching criteria and may introduce bias. However, estimates of vaccine effectiveness remained similar in the sensitivity analysis that adjusted for calendar date of RT-PCR sample collection.

In summary, we found that a two-dose schedule of CoronaVac was effective in preventing symptomatic COVID-19 and more severe clinical outcomes among elderly individuals and in a setting with extensive Gamma variant transmission. However, the delayed onset of vaccine-mediated protection underscores the need to prioritise vaccine supplies and maximise the number of individuals who complete the two-dose schedule, when CoronaVac is used as part of a mass vaccination campaign that is implemented in response to a COVID-19 epidemic.

## Author contributions

All authors conceived the study. OTR, MDTH and MD completed analyses with guidance from JRA, DATC, AIK, and JC. MSST, OFPP, OTR and MDTH curated and validated the data. OTR and MDTH wrote the first draft of the manuscript. TLD, RCP, OFPP, EFMV, MA, RS, JCG, WNA provided supervision. All authors contributed to, and approved, the final manuscript. JC is the guarantor. The corresponding author attests that all listed authors meet authorship criteria and that no others meeting the criteria have been omitted.

## Ethics approval

The study was approved by the Ethical Committee for Research of Federal University of Mato Grosso do Sul (CAAE: 43289221.5.0000.0021).

## Data sharing

Deidentified databases as well as the R codes will be deposited in the repository https://github.com/juliocroda/VebraCOVID-19

## Public and Patient Involvement

Members of the public or patients were not involved in setting the research question or the outcome measures, nor were they involved in developing plans for the design of the study. No patients were asked to advise on interpretation or writing up of results.

## Transparency statement

The lead author affirms that the manuscript is an honest, accurate, and transparent account of the study being reported; that no important aspects of the study have been omitted; and that any discrepancies from the study as originally planned have been explained.

## Dissemination declaration

Results will be disseminated to the public in Manaus and across Brazil. It is not possible to disseminate results to individuals who were selected into the study due to anonymisation of the data.

## Acknowledgements

We are grateful for the Pan American Health Organization’s support and the São Paulo State in making the databases available for analysis. JC is supported by the Oswaldo Cruz Foundation (Edital Covid-19 – resposta rápida: 48111668950485). OTR is funded by a Sara Borrell fellowship (CD19/00110) from the Instituto de Salud Carlos III. OTR acknowledges support from the Spanish Ministry of Science and Innovation through the Centro de Excelencia Severo Ochoa 2019-2023 Program and from the Generalitat de Catalunya through the CERCA Program.

## Supplementary appendix

**Supplement to:** Effectiveness of the CoronaVac vaccine in the elderly population during a Gamma variant-associated epidemic of COVID-19 in Brazil: A test-negative case-control study

**Supplementary Figure 1.**
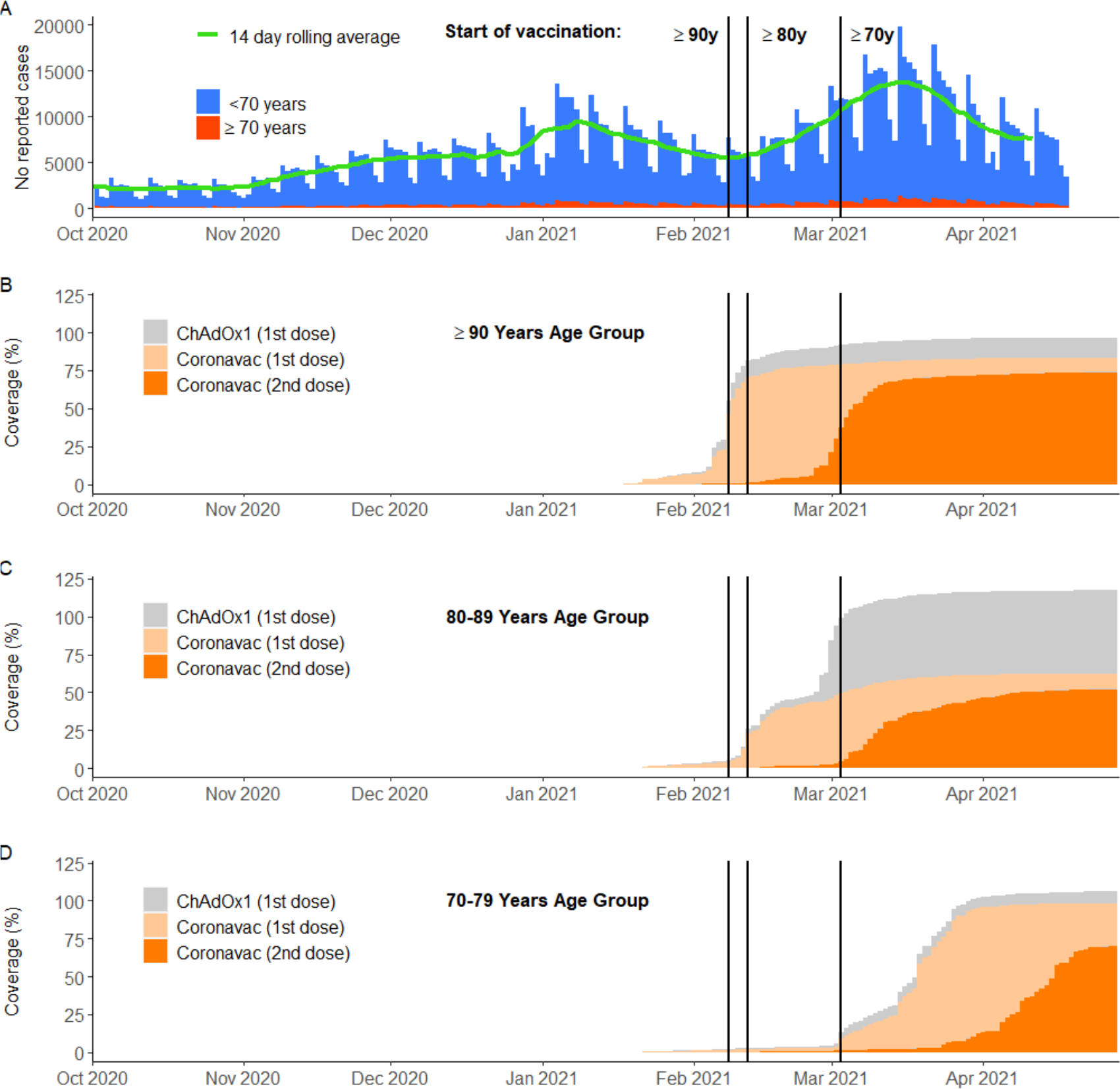
Daily cases and vaccine coverage by age. Panel A shows the daily cases of reported COVID-19 from Mar 15, 2020 to Apr 29, 2021 in São Paulo State, Brazil, with the green line representing the 14-day rolling average of counts. Panels B, C and D show the cumulative vaccination coverage for age groups >90y, 80y-89y, and 70y-79y, respectively. Population estimates for age groups were obtained from national projections for 2020.^20^ Vertical bars, from left to right in each panel, show the dates that adults ≥90, 80-89 and 70-79 years of age in the general population became eligible for vaccination.

**Supplementary Figure 2.**
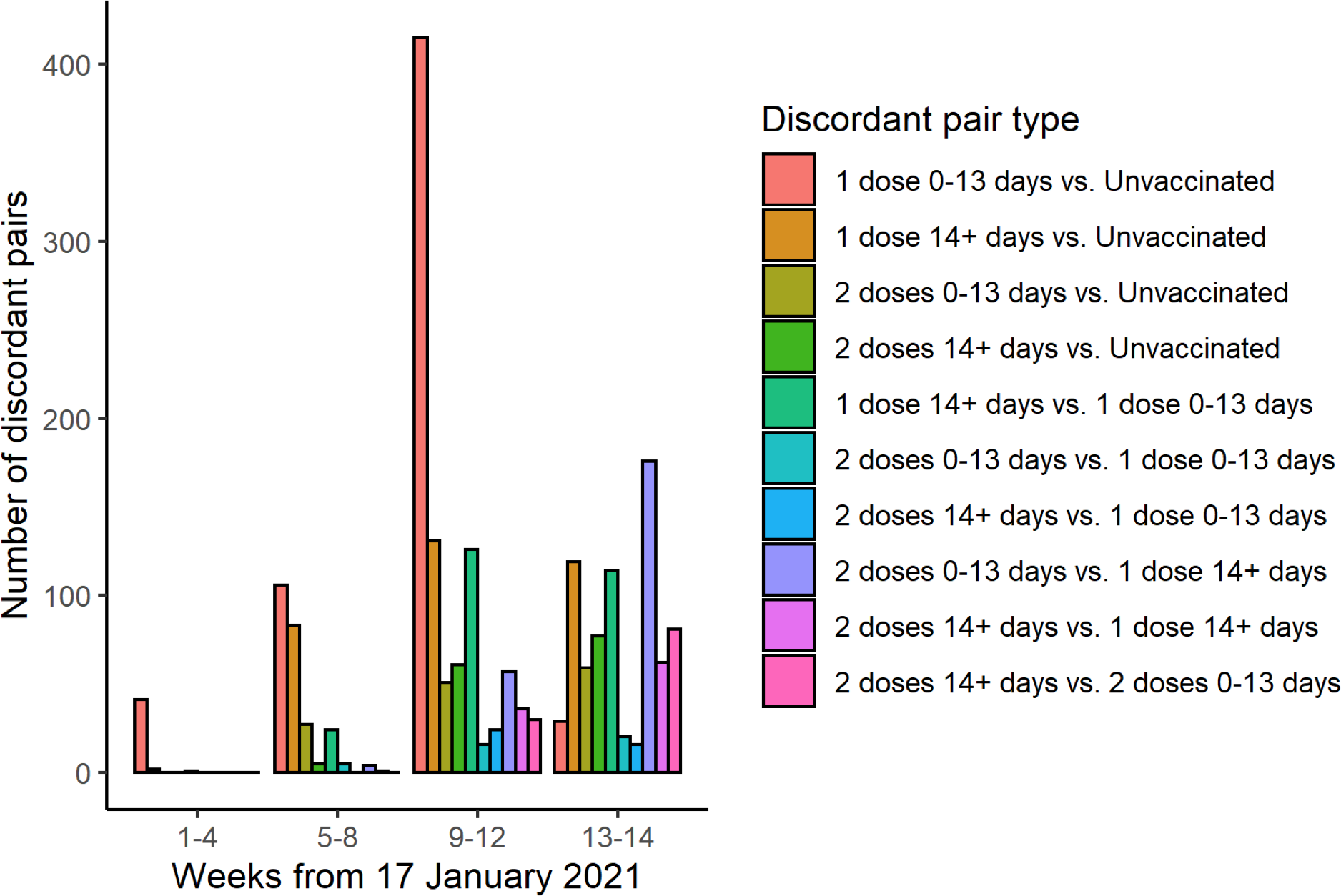
Timing of enrolment of discordant case-control pairs by vaccination category

**Supplementary Figure 3.**
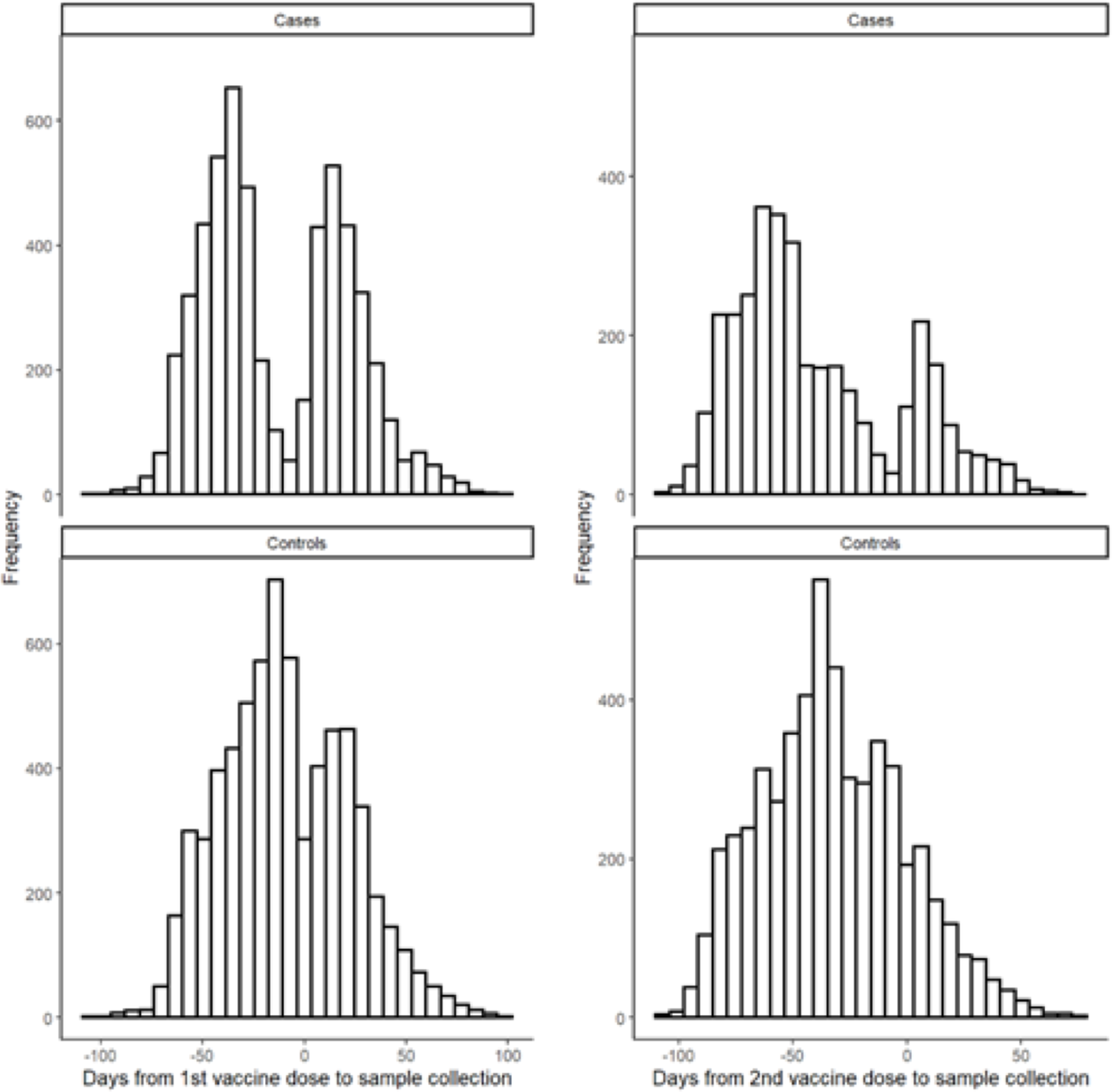
Timing of RT-PCR sample collection date relative to first (left column) and second (right column) vaccine dose date, among cases (top row) and controls (bottom row) who were vaccinated during the study period.

**Supplementary Figure 4.**
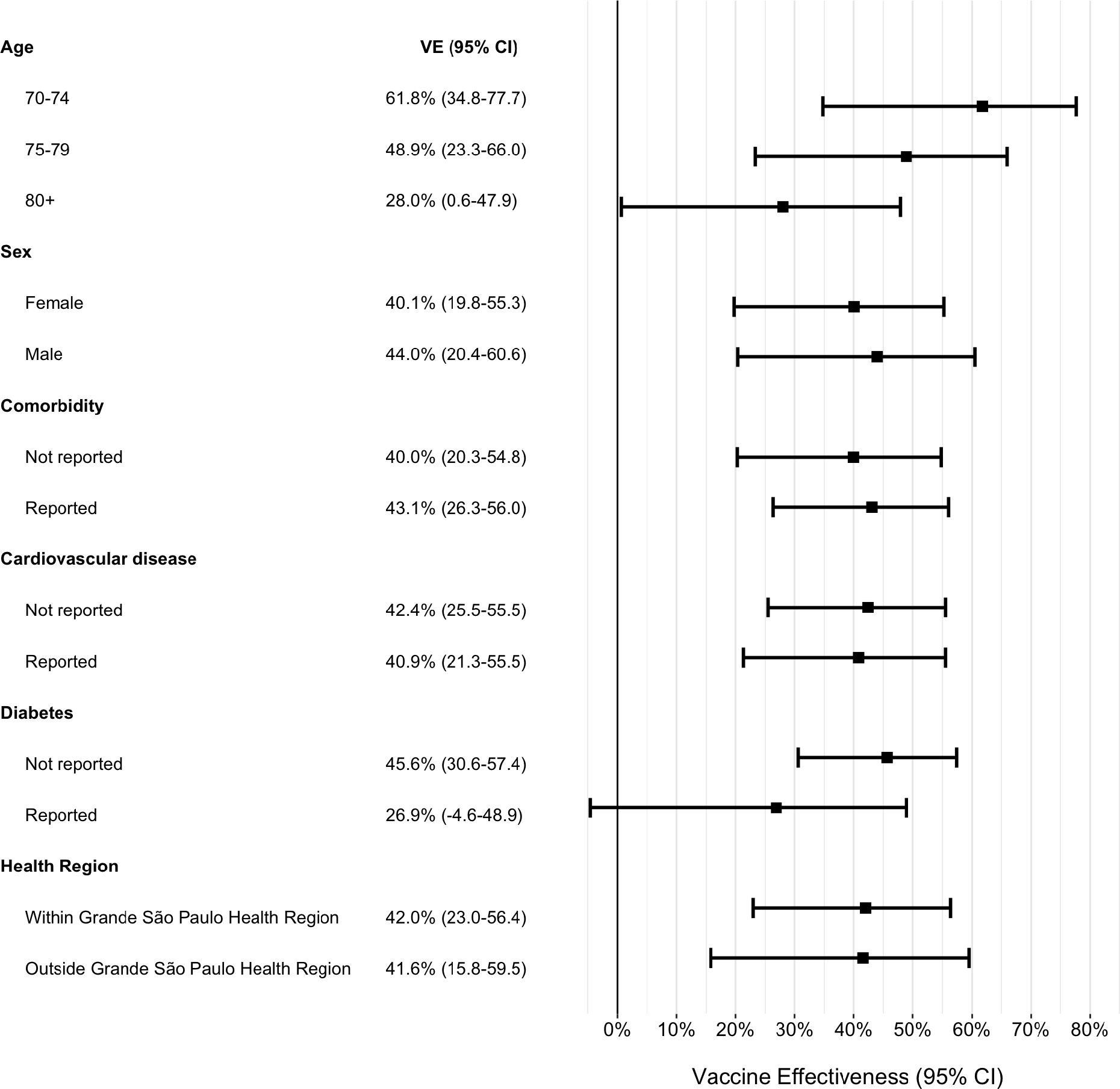
Adjusted vaccine effectiveness during the period ≥14 days after the second CoronaVac dose for subgroups of adults ≥70 years of age. Estimates of vaccine effectiveness were obtained from a conditional logistic regression model that included covariates of age (continuous) and the number of comorbidities and incorporated an interaction term between the category of interest and the period ≥14 days after the second CoronaVac dose.

**Supplementary Table 1.**
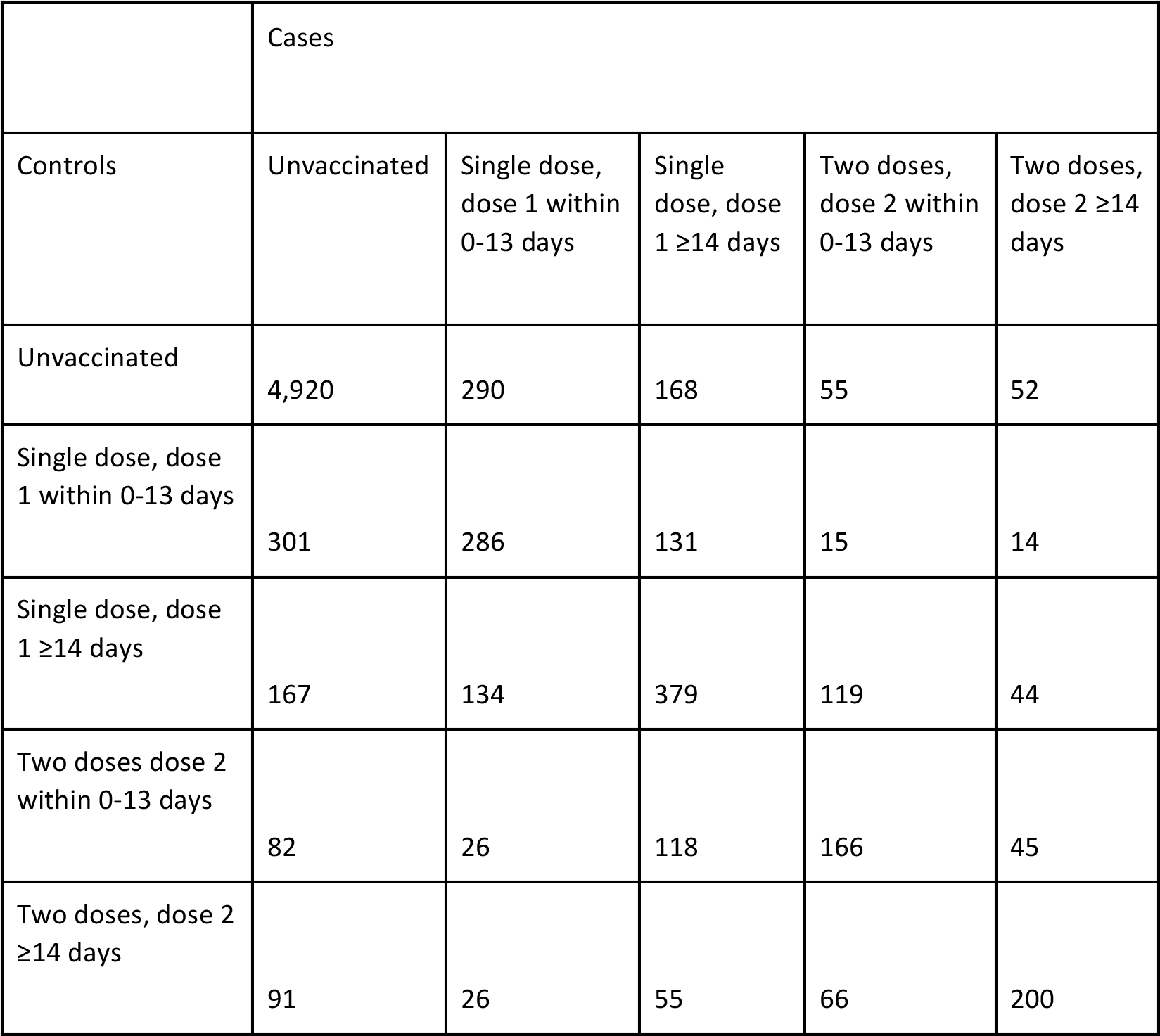
Distribution of concordant and discordant matched case-control pairs.

|  | Cases |  |  |  |  |
| --- | --- | --- | --- | --- | --- |
| Controls | Unvaccinated | Single dose, dose 1 within 0-13 days | Single dose, dose 1 $\geq 14$ days | Two doses, dose 2 within 0-13 days | Two doses, dose 2 $\geq 14$ days |
| Unvaccinated | 4,920 | 290 | 168 | 55 | 52 |
| Single dose, dose 1 within 0-13 days | 301 | 286 | 131 | 15 | 14 |
| Single dose, dose 1 $\geq 14$ days | 167 | 134 | 379 | 119 | 44 |
| Two doses dose 2 within 0-13 days | 82 | 26 | 118 | 166 | 45 |
| Two doses, dose 2 $\geq 14$ days | 91 | 26 | 55 | 66 | 200 |

**Supplementary Table 2.** Characteristics of adults ≥70 years of age who were eligible for matching and selected into case-test negative pairs for the hospitalisation analysis.

| Characteristics* | Eligible cases and controls |  | Matched pairs |  |
| --- | --- | --- | --- | --- |
|  | Test-negative<br>(n=17,622)^ | Test-positive<br>(n=26,433)^ | Controls<br>(n=4,039)^ | Cases<br>(n=4,039)^ |
| **Demographics** |  |  |  |  |
| Age, mean (SD), years | 77.53 (6.78) | 76.71 (6.19) | 77.22 (6.41) | 77.25 (6.38) |
| Age categories, n (%) |  |  |  |  |
| 70-79 years | 12,123 (68.8) | 19,673 (74.4) | 2847 (70.5) | 2847 (70.5) |
| 80-89 years | 4,301 (24.4) | 5,437 (20.6) | 965 (23.9) | 965 (23.9) |
| $\geq 90$ years | 1,198 (6.8) | 1,323 (5.0) | 227 (5.6) | 227 (5.6) |
| Male sex, n (%) | 7,689 (43.6) | 12,431 (47.0) | 1771 (43.8) | 1771 (43.8) |
| Self-reported race<sup>†</sup>, n (%) |  |  |  |  |
| White/Branca | 13,415 (76.1) | 19,796 (74.9) | 3251 (80.5) | 3251 (80.5) |
| Brown/Pardo | 3,192 (18.1) | 4,983 (18.9) | 644 (15.9) | 644 (15.9) |
| Black/Preta | 785 (4.5) | 1,258 (4.8) | 115 (2.8) | 115 (2.8) |
| Yellow/ Amarela | 226 (1.3) | 390 (1.5) | 29 (0.7) | 29 (0.7) |
| Indigenous/Indigena | 4 (0.0) | 6 (0.0) | - | - |
| Residence in “Grande São Paulo”<br>Health Region, n (%) | 12,381 (70.3) | 16,538 (62.6) | 1783 (44.1) | 1783 (44.1) |
| **Comorbidities** |  |  |  |  |
| Reported number<sup>‡</sup>, n (%) |  |  |  |  |
| None | 10,027 (56.9) | 12,668 (47.9) | 2213 (54.8) | 1127 (27.9) |
| One or two | 6,984 (39.6) | 12,548 (47.5) | 1661 (41.1) | 2566 (63.5) |
| Three or more | 611 (3.5) | 1,217 (4.6) | 165 (4.1) | 346 (8.6) |
| Cardiovascular disease , n (%) | 5,293 (30.0) | 10,079 (38.1) | 1241 (30.7) | 2201 (54.5) |
| Diabetes, n (%) | 3,233 (18.3) | 6,533 (24.7) | 793 (19.6) | 1439 (35.6) |
| **Prior SARS-CoV-2 exposure \*\*** |  |  |  |  |
| Previous symptomatic events notified to the surveillance systems**, n (%) | 685 (3.9) | 354 (1.3) | 13 (0.3) | 13 (0.3) |
| Positive SARS-CoV-2 test result <sup>††</sup>, n (%) | 66 (0.4) | 13 (0.0) | 0 (0.0) | 2 (0.0) |
| Interval between symptoms onset and RT-PCR testing, median (p25-p75), days | 3 [2-5] | 4 [2-6] | 3 [1-5] | 4 [2-6] |
| **ARI associated hospitalisations, n (%)** | 4,524/17,484 (25.9) | 12,987/26,221 (49.5) | 1,252/4,009 (31.2) | 4,039/4,039 (100) |
| **ARI associated deaths, n (%)** | 912/16,710 (5.5%) | 7,054/24,508 (28.8%) | 446/3,795 (11.8) | 1,939/3,470 (55.9) |
| Interval between symptoms onset and hospitalization, median (p25-p75), days | 3 [2-6] | 7 [4-10] | 3 [2-6] | 7 [4-10] |
| Interval between symptoms onset and deaths, median (p25-p75), days | 8 [4-13] | 14 [9-21] | 8 [4-15] | 15 [10-23] |
| **Vaccination status** |  |  |  |  |
| Not vaccinated, n (%) | 11,986 (68.0) | 17,233 (65.2) | 2656 (65.8) | 2746 (68.0) |
| Single dose, within 0-13 days, n (%) | 1,446 (8.2) | 2,976 (11.3) | 413 (10.2) | 408 (10.1) |
| Single dose, ≥14 days, n (%) | 1,797 (10.2) | 3,312 (12.5) | 445 (11.0) | 463 (11.5) |
| Two doses, within 0-13 days, n (%) | 1,041 (5.9) | 1,533 (5.8) | 230 (5.7) | 196 (4.9) |
| Two doses, ≥14 days, n (%) | 1,352 (7.7) | 1,379 (5.2) | 295 (7.3) | 226 (5.6) |
| Interval between first and second dose, mean (SD), days | 25 (6) | 30 (12) | 25 (6) | 29 (12) |
| Interval between first dose and RT-PCR testing, mean (SD), days | 28 (19) | 23 (16) | 25 (19) | 24 (18) |
| Interval between second dose and RT-PCR testing, mean (SD), days | 20 (15) | 17 (14) | 20 (16) | 20 (16) |

\*Continuous variables are displayed as mean (SD); categorical variables are displayed as n (%).
<sup>^</sup>These numbers refer to RT-PCR tests and represent 43,774 individuals for the eligible cases and controls and 8,059 individuals in the matched cases and controls.
<sup>†</sup>Race/skin colour as defined by the Brazilian national census bureau (Instituto Nacional de Geografia e Estatísticas).
<sup>‡</sup>Comorbidities included: cardiovascular, renal, neurological, haematological, or hepatic comorbidities, diabetes, chronic respiratory disorder, obesity, or immunosuppression.
<sup>\*\*</sup>Prior to the start of the study on 17 January, 2021 and after systematic surveillance was implemented on 1 February, 2020.
<sup>\*\*</sup> Reported illness with COVID-19 associated symptoms in the eSUS and SIVEP-Gripe databases.
<sup>††</sup> Defined as a positive SARS-CoV-2 RT-PCR or antigen detection test result

**Supplementary Table 3.**
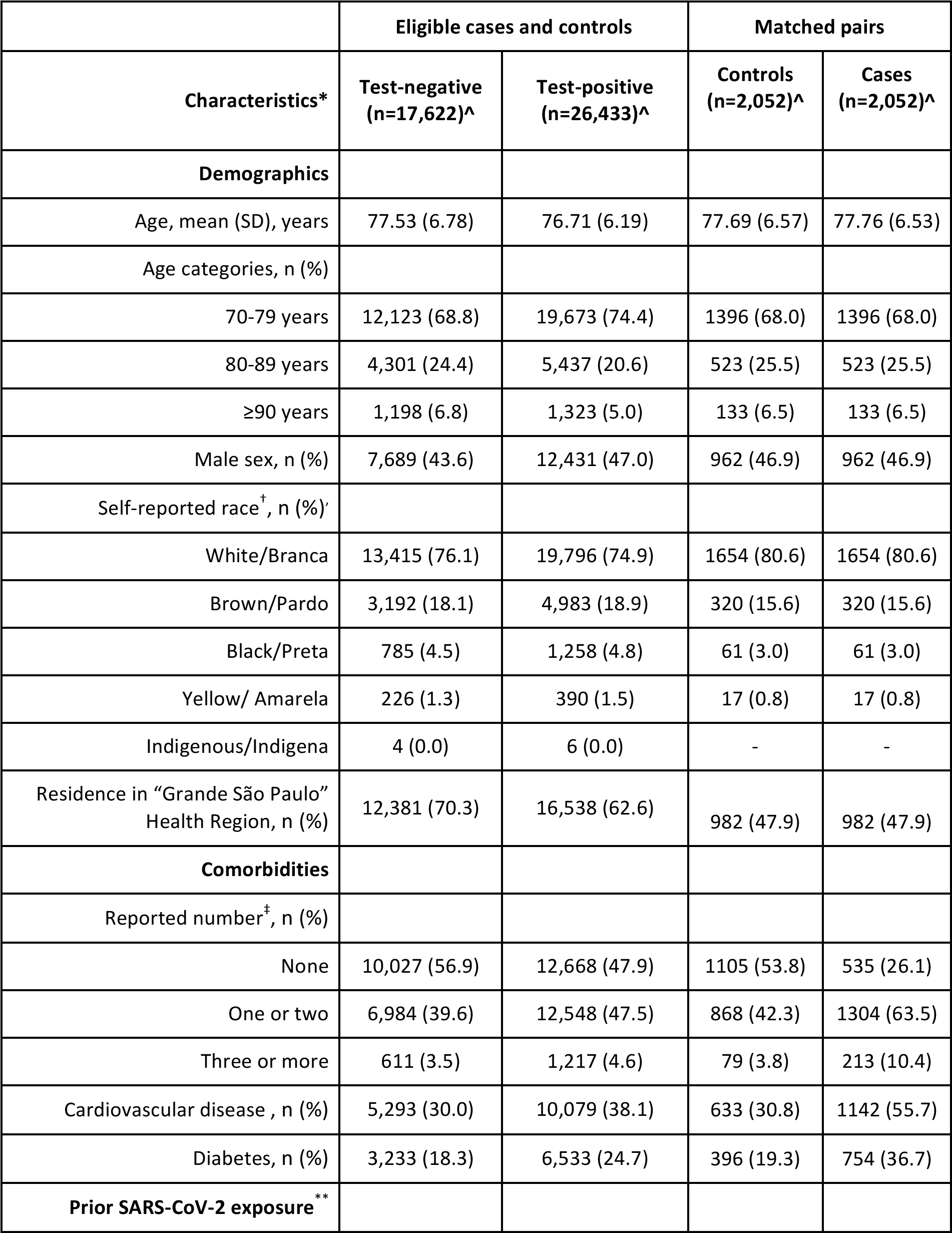
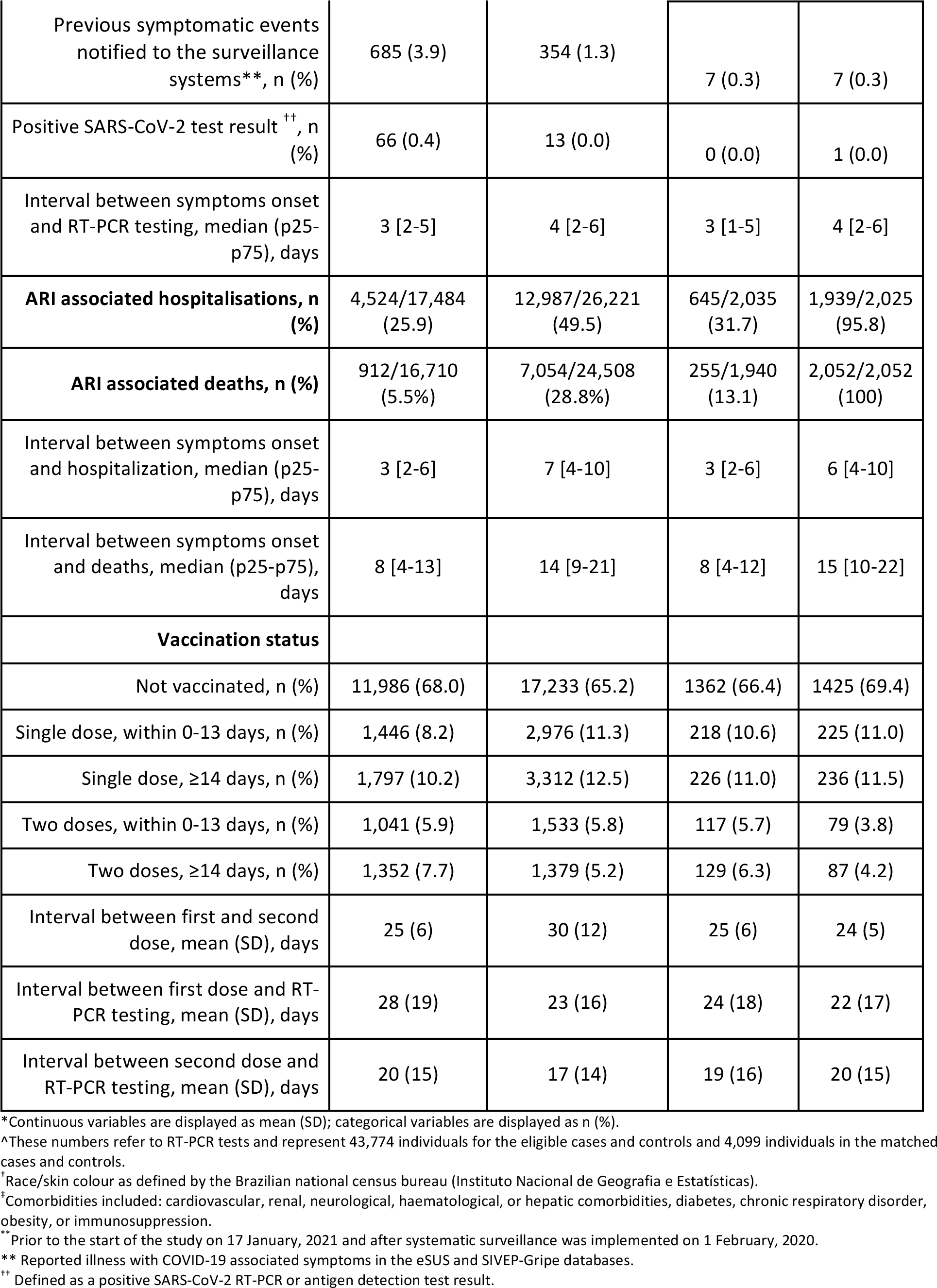
Characteristics of adults ≥70 years of age who were eligible for matching and selected into case-test negative pairs for the death analysis.

| Characteristics* | Eligible cases and controls |  | Matched pairs |  |
| --- | --- | --- | --- | --- |
|  | Test-negative<br>(n=17,622)^ | Test-positive<br>(n=26,433)^ | Controls<br>(n=2,052)^ | Cases<br>(n=2,052)^ |
| **Demographics** |  |  |  |  |
| Age, mean (SD), years | 77.53 (6.78) | 76.71 (6.19) | 77.69 (6.57) | 77.76 (6.53) |
| Age categories, n (%) |  |  |  |  |
| 70-79 years | 12,123 (68.8) | 19,673 (74.4) | 1396 (68.0) | 1396 (68.0) |
| 80-89 years | 4,301 (24.4) | 5,437 (20.6) | 523 (25.5) | 523 (25.5) |
| $\geq 90$ years | 1,198 (6.8) | 1,323 (5.0) | 133 (6.5) | 133 (6.5) |
| Male sex, n (%) | 7,689 (43.6) | 12,431 (47.0) | 962 (46.9) | 962 (46.9) |
| Self-reported race<sup>†</sup>, n (%) |  |  |  |  |
| White/Branca | 13,415 (76.1) | 19,796 (74.9) | 1654 (80.6) | 1654 (80.6) |
| Brown/Pardo | 3,192 (18.1) | 4,983 (18.9) | 320 (15.6) | 320 (15.6) |
| Black/Preta | 785 (4.5) | 1,258 (4.8) | 61 (3.0) | 61 (3.0) |
| Yellow/ Amarela | 226 (1.3) | 390 (1.5) | 17 (0.8) | 17 (0.8) |
| Indigenous/Indigena | 4 (0.0) | 6 (0.0) | - | - |
| Residence in “Grande São Paulo”<br>Health Region, n (%) | 12,381 (70.3) | 16,538 (62.6) | 982 (47.9) | 982 (47.9) |
| **Comorbidities** |  |  |  |  |
| Reported number<sup>‡</sup>, n (%) |  |  |  |  |
| None | 10,027 (56.9) | 12,668 (47.9) | 1105 (53.8) | 535 (26.1) |
| One or two | 6,984 (39.6) | 12,548 (47.5) | 868 (42.3) | 1304 (63.5) |
| Three or more | 611 (3.5) | 1,217 (4.6) | 79 (3.8) | 213 (10.4) |
| Cardiovascular disease , n (%) | 5,293 (30.0) | 10,079 (38.1) | 633 (30.8) | 1142 (55.7) |
| Diabetes, n (%) | 3,233 (18.3) | 6,533 (24.7) | 396 (19.3) | 754 (36.7) |
| **Prior SARS-CoV-2 exposure \*\*** |  |  |  |  |
| Previous symptomatic events notified to the surveillance systems**, n (%) | 685 (3.9) | 354 (1.3) | 7 (0.3) | 7 (0.3) |
| Positive SARS-CoV-2 test result <sup>††</sup>, n (%) | 66 (0.4) | 13 (0.0) | 0 (0.0) | 1 (0.0) |
| Interval between symptoms onset and RT-PCR testing, median (p25-p75), days | 3 [2-5] | 4 [2-6] | 3 [1-5] | 4 [2-6] |
| **ARI associated hospitalisations, n (%)** | 4,524/17,484 (25.9) | 12,987/26,221 (49.5) | 645/2,035 (31.7) | 1,939/2,025 (95.8) |
| **ARI associated deaths, n (%)** | 912/16,710 (5.5%) | 7,054/24,508 (28.8%) | 255/1,940 (13.1) | 2,052/2,052 (100) |
| Interval between symptoms onset and hospitalization, median (p25-p75), days | 3 [2-6] | 7 [4-10] | 3 [2-6] | 6 [4-10] |
| Interval between symptoms onset and deaths, median (p25-p75), days | 8 [4-13] | 14 [9-21] | 8 [4-12] | 15 [10-22] |
| **Vaccination status** |  |  |  |  |
| Not vaccinated, n (%) | 11,986 (68.0) | 17,233 (65.2) | 1362 (66.4) | 1425 (69.4) |
| Single dose, within 0-13 days, n (%) | 1,446 (8.2) | 2,976 (11.3) | 218 (10.6) | 225 (11.0) |
| Single dose, ≥14 days, n (%) | 1,797 (10.2) | 3,312 (12.5) | 226 (11.0) | 236 (11.5) |
| Two doses, within 0-13 days, n (%) | 1,041 (5.9) | 1,533 (5.8) | 117 (5.7) | 79 (3.8) |
| Two doses, ≥14 days, n (%) | 1,352 (7.7) | 1,379 (5.2) | 129 (6.3) | 87 (4.2) |
| Interval between first and second dose, mean (SD), days | 25 (6) | 30 (12) | 25 (6) | 24 (5) |
| Interval between first dose and RT-PCR testing, mean (SD), days | 28 (19) | 23 (16) | 24 (18) | 22 (17) |
| Interval between second dose and RT-PCR testing, mean (SD), days | 20 (15) | 17 (14) | 19 (16) | 20 (15) |

\*Continuous variables are displayed as mean (SD); categorical variables are displayed as n (%).
<sup>^</sup>These numbers refer to RT-PCR tests and represent 43,774 individuals for the eligible cases and controls and 4,099 individuals in the matched cases and controls.
<sup>†</sup>Race/skin colour as defined by the Brazilian national census bureau (Instituto Nacional de Geografia e Estatísticas).
<sup>‡</sup>Comorbidities included: cardiovascular, renal, neurological, haematological, or hepatic comorbidities, diabetes, chronic respiratory disorder, obesity, or immunosuppression.
<sup>\*\*</sup>Prior to the start of the study on 17 January, 2021 and after systematic surveillance was implemented on 1 February, 2020.
<sup>\*\*</sup> Reported illness with COVID-19 associated symptoms in the eSUS and SIVEP-Gripe databases.
<sup>††</sup> Defined as a positive SARS-CoV-2 RT-PCR or antigen detection test result.

**Supplementary Table 4.** Adjusted vaccine effectiveness during the period ≥14 days after the second CoronaVac dose for subgroups of adults ≥70 years of age. Estimates of vaccine effectiveness were obtained from a conditional logistic regression model that included covariates of age and the number of comorbidities and incorporated an interaction term between the category of interest and the period ≥14 days after the second CoronaVac dose.

| Outcome | OR (95% CI) | VE (95% CI) | p-value for interaction |
| --- | --- | --- | --- |
| **Symptomatic cases (n=15,900)** |  |  |  |
| 70-74 (n=8,178) | 0.38 (0.22-0.65) | 61.8% (34.8-77.7) | 0.05 |
| 75-79 (n=4,122) | 0.51 (0.34-0.77) | 48.9% (23.3-66.0) |  |
| 80+ (n=3,600) | 0.72 (0.52-0.99) | 28.0% (0.60-47.9) |  |
| **Hospitalisations (n=8,078)** |  |  |  |
| 70-74 (n=3,596) | 0.20 (0.09-0.44) | 80.1% (55.7-91.0) | 0.04 |
| 75-79 (n=2,098) | 0.31 (0.16-0.58) | 69.5% (42.4-83.8) |  |
| 80+ (n=2,384) | 0.57 (0.38-0.85) | 43.4% (15.4-62.0) |  |
| **Deaths (n=4,104)** |  |  |  |
| 70-74 (n=1,652) | 0.14 (0.04-0.50) | 86.0% (50.4-96.1) | 0.19 |
| 75-79 (n=1,140) | 0.13 (0.04-0.40) | 87.1% (60.2-95.8) |  |
| 80+ (n=1,312) | 0.50 (0.27-0.92) | 49.9% (8.1-72.7) |  |

**Supplementary Table 5.** Estimated effectiveness of CoronaVac ≥14 days after the second dose, in subgroups of adults ≥70 years of age. All models are adjusted by age (continuous) and number of comorbidities, and include an interaction term between the subgroup of interest and vaccinations with 2 doses, ≥14 days after second vaccine dose.

| Subgroup | Adjusted OR (95% CI) | Adjusted VE (95% CI) | p-value for interaction |
| --- | --- | --- | --- |
| **Age** |  |  |  |
| 70-74 (n=8,178) | 0.38 (0.22-0.65) | 61.8% (34.8-77.7) | 0.05 |
| 75-79 (n=4,122) | 0.51 (0.34-0.77) | 48.9% (23.3-66.0) |  |
| 80+ (n=3,600) | 0.72 (0.52-0.99) | 28.0% (0.60-47.9) |  |
| **Sex** |  |  |  |
| Females (n=9,348) | 0.60 (0.45-0.80) | 40.1% (19.8-55.3) | 0.85 |
| Males (n=6,552) | 0.56 (0.39-0.80) | 44.0% (20.4-60.6) |  |
| **Comorbidities** |  |  |  |
| No reported (n=8,074) | 0.60 (0.45-0.80) | 40.0% (20.3-54.8) | 0.81 |
| Reported (n=7,826) | 0.57 (0.44-0.74) | 43.1% (26.3-56.0) |  |
| **Cardiovascular disease** |  |  |  |
| No reported (n=10,273) | 0.58 (0.45-0.75) | 42.4% (25.5-55.5) | 0.86 |
| Reported (n=5,627) | 0.59 (0.45-0.79) | 40.9% (21.3-55.5) |  |
| **Diabetes** |  |  |  |
| No reported (n=12,294) | 0.54 (0.43-0.69) | 45.6% (30.6-57.4) | 0.12 |
| Reported (n=5,627) | 0.73 (0.51-1.05) | 26.9% (-4.6-48.9) |  |
| **Health regional area** |  |  |  |
| “Grande São Paulo” (n=7,382) | 0.58 (0.44-0.77) | 42% (23.0-56.4) | 0.66 |
| Not “Grande São Paulo” (n=8,518) | 0.58 (0.41-0.84) | 41.6% (15.8-59.5) |  |

### Protocol for the Teste-Negative Case-Control Study in São Paulo State

Released in https://github.com/juliocroda/VebraCOVID-19/

#### PROTOCOL

Evaluation of Vaccine Effectiveness in Brazil against COVID-19 (VEBRA-COVID) Sub-Study: A Test-Negative Case-Control Study on the Effectiveness of COVID-19 Vaccines amongst the General Population of São Paulo State in Brazil

**Table 1.** Protocol Revisions

| **Changes in Version 1.3** | **Justification** |
| --- | --- |
| Addition of ChAdOx1 exposure times | We added the time windows following the first and second doses of ChAdOx1 to be 0-13 days, 14-27 days and $\geq 28$ days |
| Revised expected vaccine effectiveness | In the VEBRA-COVID analysis of the elderly ($\geq 70$ years of age) in São Paulo, we aimed to answer the research question of whether vaccines had a real-world effectiveness of public health value rather than whether they had a real-world effectiveness that was consistent with efficacy estimates from RCTs. Thus, we powered the study for a real world effectiveness above a lower threshold of 40%, below which the value of the vaccination would require reconsideration. |
| Change of matching criteria from CEP (5 digits) to Municipality and self-reported race | We based this decision on three main reasons:<br>1 – A great proportion of municipalities in São Paulo State has a unique CEP (zipcode), so everyone in that municipality has the same CEP. For these municipalities, we would lose within municipality socioeconomic information<br>2 – We observed a larger proportion of invalid CEPs mainly in the e-SUS database compared with the SIVEP-Gripe database, which may introduce potential bias since SIVEP-Gripe has a higher proportion of severe COVID-19 cases<br>3 – A significant number of unique CEPs were inconsistently placed in more than one municipality. |
| Addition of outcomes for the cohort analysis of test-positive cases | We added ICU admission and respiratory support, occurring within 21 days of initial SARS-CoV-2 test positivity. We also changed hospitalization from occurring within 14 days to within 21 days of initial SARS-CoV-2 test positivity. |

##### I. Background

Since the emergence of severe acute respiratory virus coronavirus 2 (SARS-CoV-2), Brazil has experienced one of the world’s highest incidence and mortality rates in the world, with over 13 million reported infections as of the middle of April 2021.^1–3^ São Paulo, the most populous state in Brazil (∼ 46 million inhabitants), is the state with highest number of cases and deaths: 2,827,833 cases and 92,548 deaths as by April 24^th^ 2021.^4^ Variants of Concern (VOC) also had a key role on the recent several surges in Brazil and São Paulo State. The P.1 VOC, which was first detected in Manaus on Jan 12, 2021, ^5–7^ and now consists the majority of new infections, being dominant in several states in Brazil. P1. has accrued mutations associated with decreased neutralization,^8, 9^ and has since spread throughout Brazil, synchronizing the epidemic in country in a scenario of relaxed non-pharmacological interventions.

The rapid development of novel vaccines against COVID-19 allowed countries to start vaccine distribution programs within a year of the identification of the novel virus. Among the first vaccines to be developed was Sinovac’s CoronaVac vaccine.^10–12^ Phase III trials were conducted in Turkey, Chile, Singapore and Brazil. The Brazilian trial was conducted among a study population of healthcare professionals, and reported that the effectiveness of CoronaVac after 14 days following completion of a two dose schedule was 50.7% (95% CI 36.0-62.0) for all symptomatic cases of COVID-19, 83.7% (95% CI 58.0-93.7) for cases requiring medical attention, and 100% (95% CI 56.4-100) for hospitalized, severe, and fatal cases.^12^ CoronaVac was approved for emergency use on 17 January in Brazil, and used to vaccinate healthcare workers and the general population. AstraZeneca-Oxford’s ChAdOx1 vaccine^13, 14^ was approved on the same day and was administered beginning on 23 January 2021. In Brazil, ChAdOx1 schedule is for 12 weeks between first and second dose.

As vaccine programs continue, there has been much interest in estimation of vaccine effectiveness through observational studies, and specifically in settings where VOC are circulating. Such studies have advantages over clinical trials, including increased size and follow-up time, and reduced cost. However, as vaccinated and unvaccinated individuals are likely different in their SARS-CoV-2 risk and healthcare access, these studies must address bias through design and analysis. Several studies have demonstrated the effectiveness of COVID-19 vaccines against infection caused by the B.1.1.7 variant.^15^ However, large-scale real-world investigations on vaccine effectiveness have not been conducted in regions where the P.1 variant is prevalent.

We propose a test-negative case-control study^16, 17^ of the general population from the São Paulo State to evaluate the effectiveness of COVID-19 vaccines in preventing symptomatic disease in a setting of widespread P.1 VOC transmission.^6^ The study will initially evaluate the effectiveness of COVID-19 vaccines, CoronaVac and ChAdOx1 amongst the population with age ≥70 years, since the vaccination campaign prioritized this age group in its first months. We will expand the study population as additional age groups become eligible for vaccination. Furthermore, we expect that additional vaccines will be approved and will evaluate their effectiveness. We will therefore continue to amend the protocol and its objectives accordingly to address these new questions.

##### II. Objectives

To estimate the effectiveness of COVID-19 vaccines against symptomatic SARS-CoV-2 infection amongst the general population from the São Paulo State. Our initial analyses will focus on estimating vaccine effectiveness in the age group of ≥70 years.

##### III. Methods

###### 1. Study Design

We will conduct a retrospective matched case-control study, enrolling cases who test positive for SARS-CoV-2 and controls who test negative for SARS-CoV-2 amongst the general population (Section 3) as of the day that the COVID-19 vaccination campaign was initiated at the study sites. The study will evaluate vaccine effectiveness on the primary outcome of symptomatic SARS-CoV-2 infection. We will identify cases and matched controls by extracting information from health surveillance records and ascertain the type and data of vaccination by reviewing the state COVID-19 vaccination registry. In this design, one minus the odds ratio (1-OR) of vaccination comparing cases and controls estimates the direct effect of vaccination on the disease outcome. In a separate analysis, we will assess the association between vaccination and hospitalization and/or death among individuals who have tested positive for SARS-CoV-2.

###### 2. IRB and Ethics Statement

The protocol has been submitted to the Ethical Committee for Research of Federal University of Mato Grosso do Sul (CAAE: 43289221.5.0000.0021). The work of investigators at the University of Florida, Yale University, Stanford University, and Barcelona Institute for Global Health was conducted to inform the public health response and was therefore covered under Public Health Response Authorization under the US Common Rule.

###### Study Site

The State of São Paulo (23°3′S, 46°4’W) is the most populous state in Brazil: an estimated population of 46,289,333 in 2020. São Paulo State has 645 municipalities and its capital, São Paulo city, has 12 million inhabitants. São Paulo State reported 2,827,833 COVID-19 cases (cumulative incidence rate: 6,109 per 100,000 population) and 92,548 deaths (cumulative mortality: 200 per 100,000 population), by 24/04/2021. The State Secretary of Health of Sao Paulo (SES-SP) initiated its COVID-19 vaccination campaign on 17 January 2021 and is administering two vaccines, CoronaVac and ChAdOx1. As of 24 April 2021, 10.7 million doses (6.9 million first doses and 3.8 million second doses) have been administered in the State.

###### Data Sources and Integration

We will identify eligible cases and controls from the State of São Paulo who test positive and negative, respectively, from the *state laboratory testing registry* of public health laboratory network; 2) Determine vaccination status from *state vaccination registries*; and 3) Extract information from *national healthcare and surveillance databases* that will be used to define outcomes, match controls to cases, determine vaccination status, serve as covariates for post-stratification and provide a source for cross-validation of information from databases. Registries are not available which enables constructing a cohort of people eligible for vaccination in the general population. Data sources for this study will include:

- State laboratory testing registry (**GAL**) of the network of public health laboratories
- State COVID-19 vaccination registry (**Vacina Já**)
- National surveillance database of severe acute respiratory illnesses (**SIVEP-Gripe**) created by Ministry of Health Brazil in 2009
- National surveillance system of suspected cases of COVID-19 (**e-SUS**) from mild to moderate “influenza like illness”, created by the Ministry of Health Brazil in 2020

The databases will be integrated by the São Paulo State Government – PRODESP - using CPF numbers (Brazilian citizens’ unique identifier code) and send to the VEBRA-COVID group anonymized. The database will be updated on a bi-weekly basis.

###### Study Population

Inclusion criteria:

● Has a residential address in the State of São Paulo,
● Eligible to receive a COVID-19 vaccine based on age,
● With complete information, which is consistent between databases, on age, sex, and residential address
● With consistent vaccination status and dates for those who were vaccinated.

Exclusion criteria:

● Does not have a residential address in the State of São Paulo,
● Not eligible to receive a COVID-19 vaccine based on age,
● With missing or inconsistent information on age, sex, or city of residence
● With existing but inconsistent vaccination status or dates.

###### Case definition and eligibility

We will use information from integrated GAL/SIVEP-Gripe/e-SUS databases to identify cases that are defined as eligible members of the study population (as defined above, Study Population) who:

- Had a sample with a positive SARS-CoV-2 RT-PCR, which was collected between January 17, 2021 and 7 days prior to database extraction of information
- Did not have a positive RT-PCR test in the 90 day period preceding the index positive RT-PCR result
- Have complete and consistent data on SARS-CoV-2 RT-PCR test results

###### Control definition and eligibility

We will use integrated GAL/SIVEP-Gripe/e-SUS databases to identify eligible controls. Controls are defined as eligible members of the study population who:

- Had a sample with a negative SARS-CoV-2 RT-PCR result, which was collected after January 17, 2021,
- Did not have a positive RT-PCR test in the 90 day period preceding the index positive RT-PCR result
- Did not have a subsequent positive RT-PCR test in the 7-day period following the index positive RT-PCR result
- Have complete and consistent data on SARS-CoV-2 PCR test result

When studying each vaccine, individuals that received another vaccine are eligible for selection as a case and/or control until the day they receive their vaccine, i.e. we will consider test positive and test negative cases for RT-PCR collected before the day of receipt of the other vaccine.

###### Matching

Test-negative controls will be matched 1:1 to the cases. We chose the matching factors to balance the ability to reduce bias and to enroll sufficient case-control pairs. Matching factors will include variables that are anticipated to be causes of the likelihood of receiving the vaccine, risk of infection and likelihood of receiving PCR testing for SARS-CoV-2 (see Figures 1-5):

- Age, categorized as 5-years age bands (e.g., 70-74, 75-79 years),
- Sex,
- Municipality,
- Self-reported race,
- Window of ±3 days between collection of RT-PCR positive respiratory sample for cases and collection of RT-PCR negative respiratory sample for controls. If the date of respiratory sample collection is missing, the date of notification of testing result will be used.

We will use the standard algorithms to conduct matching which include: 1) setting a seed, 2) locking the database, 4) creating a unique identifier for matching after random ordering, 5) implementing exact matching based on matching variables, sampling controls at random if more than one available per case within strata.

An individual who fulfils the control definition and eligibility and later has a sample tested that fulfils the case definition and eligibility can be included in the study as both a case and a control. An individual who fulfils the control definition for multiple different sample collection dates can be included in the study as a control for each collection date, up to a maximum of three times.

##### Exposure definition

CoronaVac vaccination:

- Received the first vaccine dose, and not having received a second dose, in the following time periods relative to sample collection for their PCR test:

0-13 days
≥14 days
- Received the second dose in the following time periods relative to sample collection for their PCR test:

0-13 days
≥14 days

ChAdOx1vaccination:

- Received the first vaccine dose, and not having received a second dose, in the following time periods relative to sample collection for their PCR test:

0-13 days
14-27 days
≥28 days
- Received the second dose in the following time periods relative to sample collection for their PCR test:

0-13 days
≥14 days

##### Statistical Analyses

We will evaluate the effectiveness of CoronaVac and ChAdOx1for the following SARS-CoV-2 infection outcomes:

- Primary: Symptomatic COVID-19, defined as one or more reported COVID-19 related symptom with onset within 0-10 days before the date of their positive RT-PCR test
- Secondary:

COVID-19 associated hospitalization within 21 days of the symptom onset
COVID-19 associated ICU admission within 21 days of the symptom onset
COVID-19 associated respiratory support
COVID-19 associated death within 28 days of symptom onset

We will evaluate vaccine effectiveness for the primary outcome according to the test-negative design. Table 1 shows a list of all planned analyses in the test-negative design. The test-negative design may introduce bias when evaluating outcomes of hospitalizations and deaths during an epidemic. We will therefore perform time to event/logistic regression analysis of test positive cases to evaluate the association of vaccination status and the risk for hospitalization, ICU admission, COVID-19 respiratory support, and death after infection.

Our initial analyses will focus on estimating vaccine effectiveness in the population with age ≥70 years of age who were the initial priority group of the COVID-19 vaccination campaign.

##### Case-control analysis

Analyses of the primary outcome will be restricted to case and control pairs who are matched based on the presence of a COVID-19 related symptom before or at the time of testing.

We will use conditional logistic regression to estimate the odds ratio (OR) of vaccination among cases and controls, accounting for the matched design, where 1-OR provides an estimate of vaccine effectiveness under the standard assumptions of a test-negative design. For the CoronaVac analysis, the reference group will be individuals who have not received a first dose of CoronaVac by the date of respiratory sample collection. For the ChAdOx1 analysis, the reference group will be individuals who have not received a first dose of ChAdOx1by the date of respiratory sample collection. Date of notification of the testing result will be used if the date of respiratory sample collection is missing. To evaluate potential biases and the timing of vaccine effectiveness after administration, we will evaluate the windows of vaccination status corresponding: A) 0-13 days and ≥14 days after the 1^st^ dose and 0-13 days and ≥14 days after the 2^nd^ dose of CoronaVac; and B) 0-13 days, 14-27 days and ≥28 after the 1st dose and0-13 days and ≥14 days after the 2^nd^ dose of ChAdOx1.

We will include the following covariates in the adjusted model, which we hypothesize are predictive of vaccination, the risk of SARS-CoV-2 infection and COVID-19 severity and healthcare access and utilization:

- Age as continuous variable
- Comorbidities (None, 1-2, ≥3 comorbidities)
- Evidence of prior SARS-CoV-2 infection (defined as positive PCR test, antigen test or rapid antibody test)

Although data on comorbidities is available through e-SUS and SIVEP-Gripe, this data may have different degrees of missingness between databases and between cases and control groups. Adjusting for comorbidities using complete case data will likely introduce bias. We will explore the feasibility of multiple imputation of comorbidity in a sensitivity analysis. Additional sensitivity analyses will evaluate potential effect modification of the vaccine effectiveness by history of a positive RT-PCR, antigen or serological test result prior to the vaccination campaign and age subgroups.

##### Survival/logistic regression analysis of hospitalization, ICU, respiratory support and death

We will perform additional analyses for hospitalization and death amongst individuals who test positive and estimate the hazards according to vaccination status at the date of positive test, adjusting for covariates described in the case-control analyses. Sensitivity analyses will be conducted to evaluate the association of influence of a positive RT-PCR, antigen or serological test result prior to the vaccination campaign.

##### Sample size calculations and timing of analyses

The power of a matched case-control study depends on the assumed odds ratio and the number of discordant pairs (i.e. pairs in which the case is exposed and the control is unexposed, or vice versa), which is a function of the assumed odds ratio and the expected prevalence of exposure among controls. Moreover, the estimate of the odds ratio for one level of a categorical variable compared to baseline is determined by the distribution of all discordant pairs. As vaccine coverage and incidence are changing over time, the latter in ways we cannot predict, and there is no power formula for this analysis, we will simulate power and enroll individuals until we have reached a target power, which we can assess without analyzing the data. In particular, after determining the number of discordant case-control pairs for each combination of exposure categories, we will randomly assign one of each pair to each relevant exposure type according to a Bernoulli distribution, with the probability determined by the assumed odds ratio comparing the two categories. We will run an unadjusted conditional logistic regression on the simulated dataset to determine the p-value, and estimate the power as the proportion of N=1,000 simulations that return p<0.05. Code to perform the power calculation can be found at https://github.com/mhitchings/VEBRA_COVID-19.

##### Timing of final analyses

We will perform an analysis of the primary outcome upon reaching simulated 80% power to detect vaccine effectiveness of 40% ≥14 days after the second dose for the CoronaVac. For the ChAdOx1, we will perform an analysis of effectiveness of at least one dose upon reaching simulated 80% power to detect vaccine effectiveness of 40% ≥28 days after the first dose. In addition, we will perform an analysis of effectiveness of two doses upon reaching simulated 80% power to detect vaccine effectiveness of 40% ≥14 days after the second dose. We chose a vaccine effectiveness of 40% to address the question of whether vaccination with CoronaVac and ChAdOx achieved a threshold of real-world effectiveness, below which the public health value of vaccination may need to be reconsidered.

##### Privacy

Only SES-SP, São Paulo State data management had access to the identified dataset to linkage the datasets by name, date of birth, mother’s name and CPF. After the linkage, the CPF was encrypted and the de-identified dataset was sent to the team for analysis.

##### Working group

Matt Hitchings, Otavio T. Ranzani, Julio Croda, Albert I. Ko, Derek Adam Cummings, Wildo Navegantes de Araujo, Jason R. Andrews, Roberto Dias de Oliveira, Patricia Vieira da Silva, Mario Sergio Sacaramuzzini Torres, Wade Schulz, Tatiana Lang D Agostini, Edlaine Faria de Moura Villela, Regiane A. Cardoso de Paulo, Olivia Ferreira Pereira de Paula, Jean Carlo Gorinchteyn

**Figure 1:**
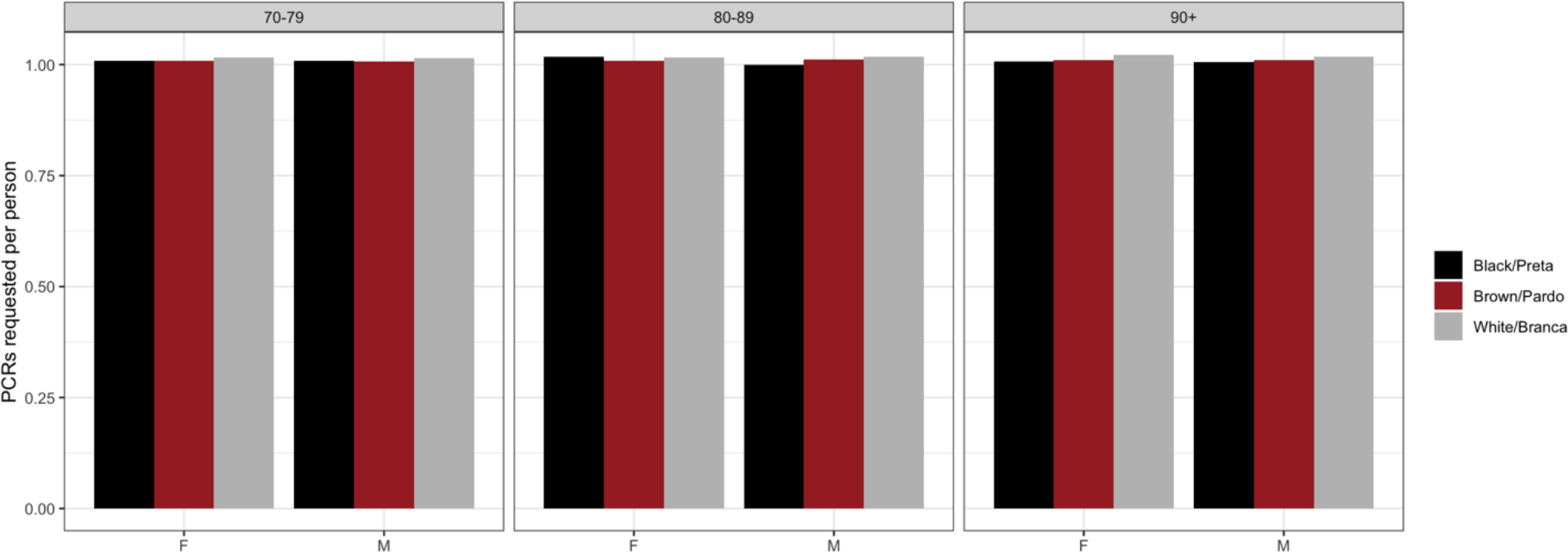
PCR testing rate by age, sex and self-reported race (from data extracted on April 07, 2021)

**Figure 2:**
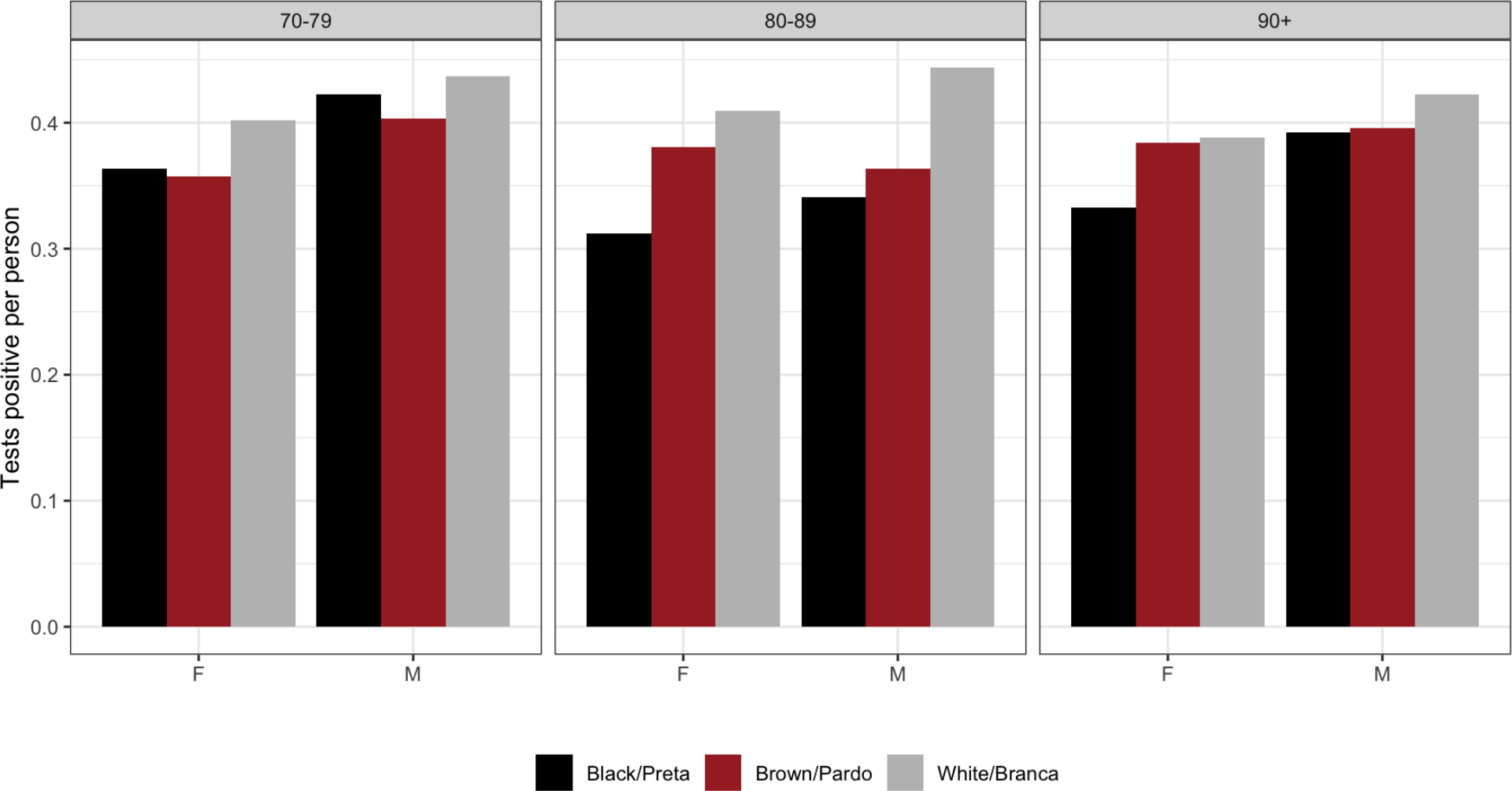
PCR positive testing rate by age, sex and self-reported race (from data extracted on April 07, 2021)

**Figure 3:**
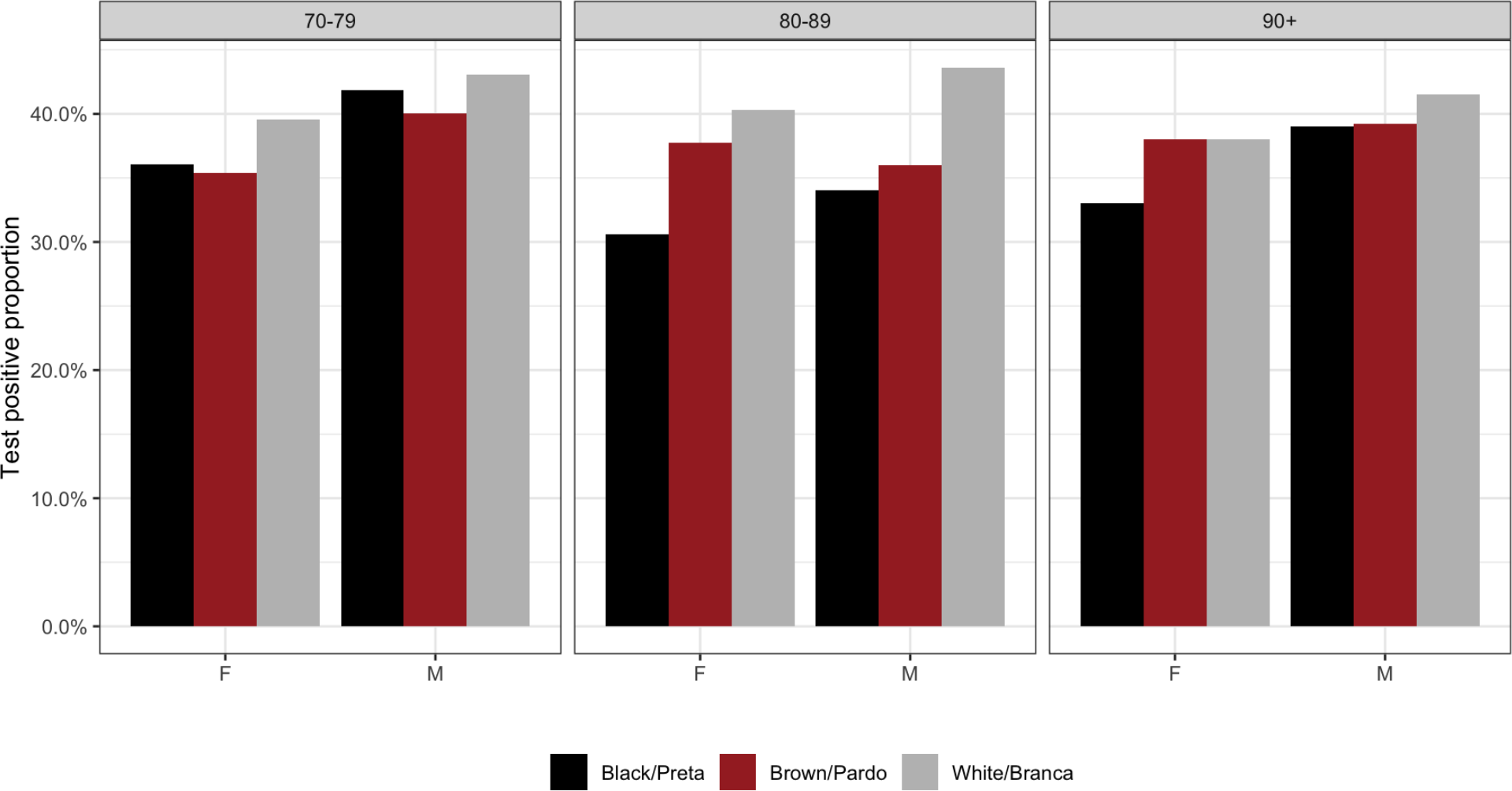
PCR positive proportion by age, sex and self-reported race (from data extracted on April 07, 2021)

**Figure 4:**
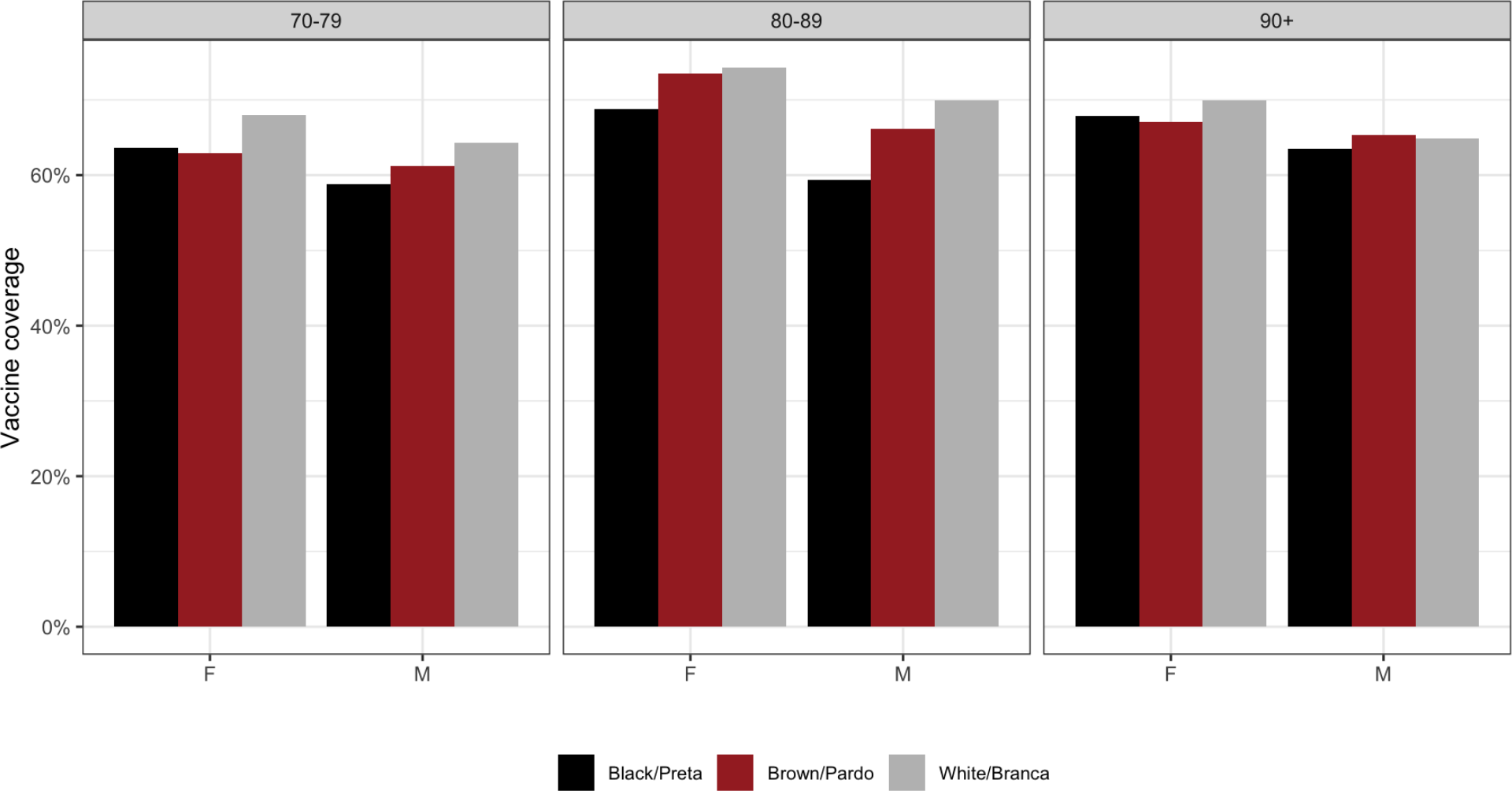
Vaccine coverage by age, sex and self-reported race (from data extracted on April 07, 2021)

**Figure 5:**
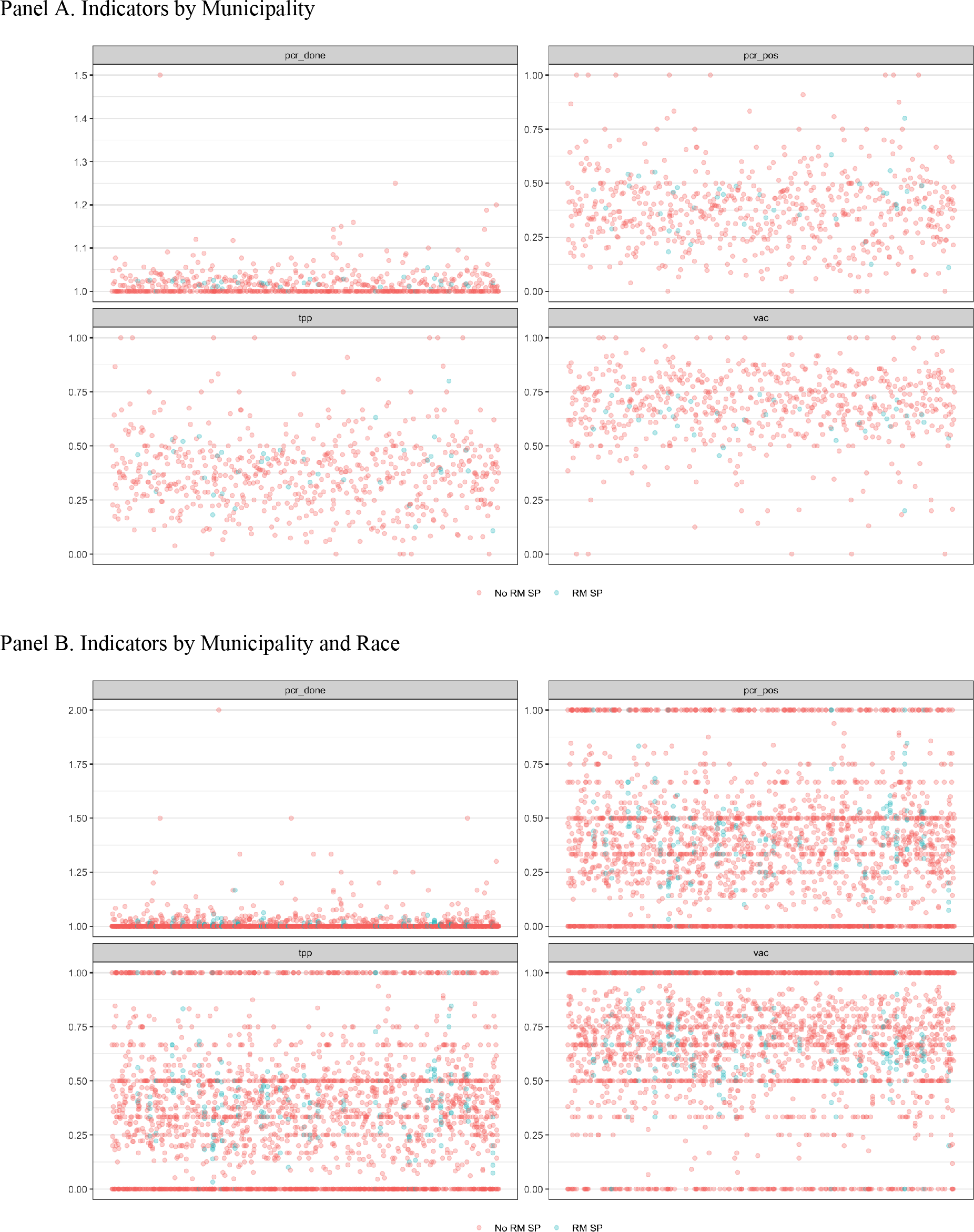
PCR testing rate (pcr_done), PCR positive testing rate (pcr_pos), positivity proportion (tpp) and vaccine coverage (vac) by each municipality (A) and municipality and race (B). RM SP denotes metropolitan area of São Paulo city (from data extracted on April 07, 2021)

**Supplementary Figure 1.**
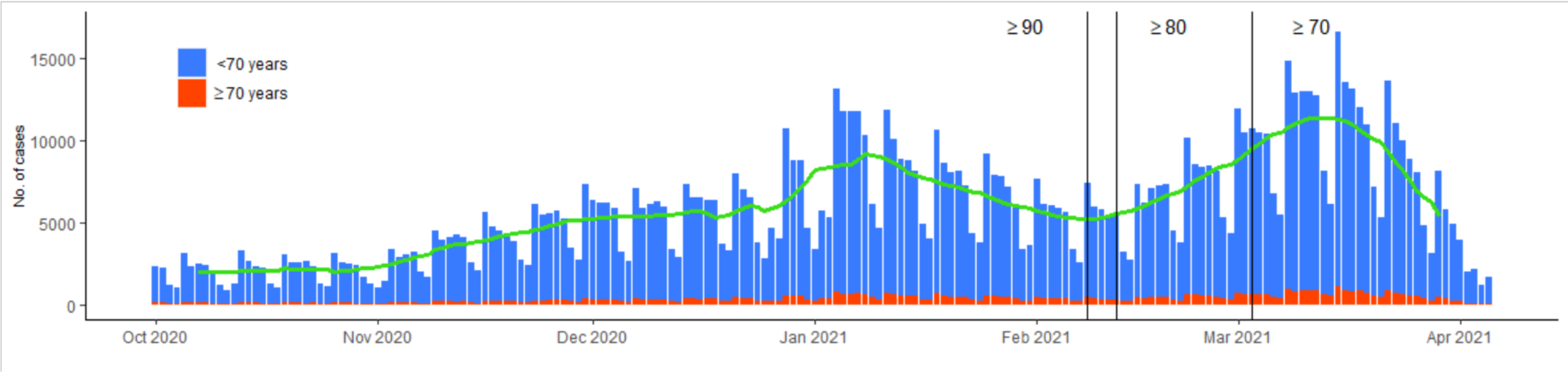
**Reported RT-PCR or Antigen confirmed COVID-19 in the general population of the São Paulo State, Brazil from October 2020 to April 7, 2021. Lines depict moving 14-day averages for case. Vertical lines represent vaccine eligibility by age.**

**Supplementary Figure 2.**
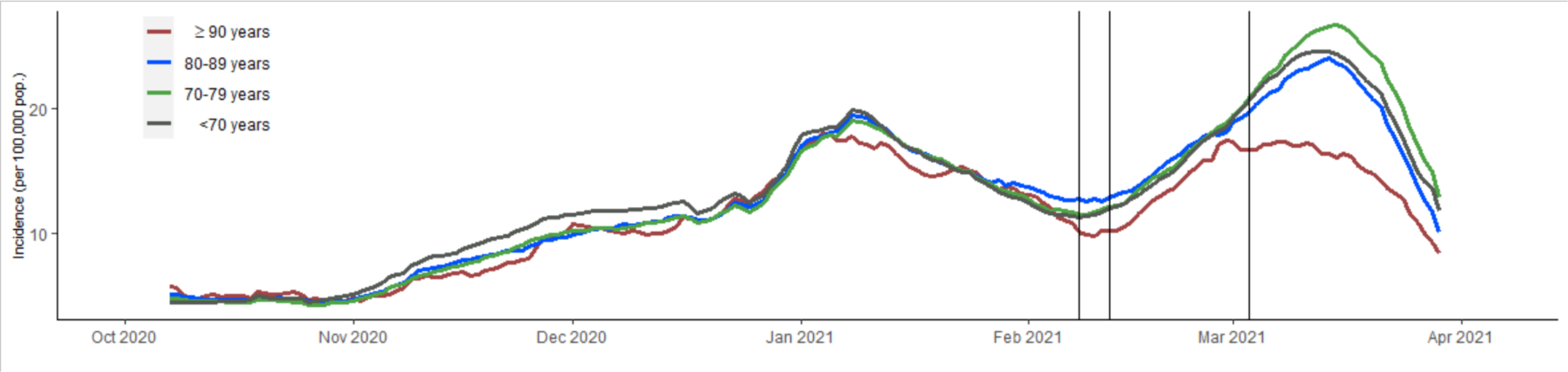
**Reported RT-PCR or Antigen confirmed COVID-19 rates in the general population of the São Paulo State, Brazil from October 2020 to April 7, 2021. Lines depict rolling averages. Vertical lines represent vaccine eligibility by age.**

**Table 1.** Table of planned

| Analysis | Exposure | Outcome |
| --- | --- | --- |
| **CoronaVac** |  |  |
| Primary outcome, primary exposure | Two-dose regimen of CoronaVac in the period starting 14 days after administration of the 2<sup>nd</sup> dose | Positive test for SARS-CoV-2, with at least one COVID-19 symptom reported 0-10 days before sample collection date |
| Primary outcome, secondary exposure (2-dose) | Two-dose regimen of CoronaVac in the period 0-13 days after administration of the 2<sup>nd</sup> dose |  |
| Primary outcome, secondary exposure (1-dose) | One-dose regimen of CoronaVac, in the period starting 14 days after administration of the 1<sup>st</sup> dose |  |
| Primary outcome, bias indicator | One-dose regimen of CoronaVac, in the period 0-13 days after administration of the 1<sup>st</sup> dose |  |
| **ChAdOx1** |  |  |
| Primary outcome, primary exposure | One-dose regimen of ChAdOx1 in the period starting 28 days after administration of the 1<sup>st</sup> dose | Positive test for SARS-CoV-2, with at least one COVID-19 symptom reported 0-10 days before sample collection date |
| Primary outcome, secondary exposure (2-dose) | Two-dose regimen of ChAdOx1 in the period $\geq 14$ days after administration of the 2<sup>nd</sup> dose | |
| Primary outcome, secondary exposure (1-dose) | One-dose regimen of ChAdOx1 in the period 0-13 days after administration of the 1<sup>st</sup> dose |  |
| Primary outcome, secondary exposure (1-dose) | One-dose regimen of ChAdOx1, in the period starting 14-27 days after administration of the 1<sup>st</sup> dose |  |
| Primary outcome, secondary exposure (2-dose) | Two-dose regimen of ChAdOx1, in the period starting 0-13 days after administration of the 2<sup>nd</sup> dose |  |
| Primary outcome, bias indicator | One-dose regimen of ChAdOx1, in the period 0-13 days after administration of the 1<sup>st</sup> dose |  |

